# Circulating immune architecture across age, exposures and disease in ethnically diverse UK cohorts

**DOI:** 10.64898/2026.11.17.26367600

**Authors:** Daniela Zanotti, Giuditta Clerici, Stephen Watt, Olli Dufva, Edoardo Giacopuzzi, Manuel Tardáguila Sancho, Alessandro Raveane, Arianna Landini, Jan P. Engelmann, Matiss Ozols, Paola Benaglio, Beatrice Costa, Bruno Ariano, Sodbo Sharapov, Lucas Arnoldt, Danai Vagiaki, Davide Bolognini, Eva Holtkamp, Shuang Li, Ruth Eberhardt, Andrea Senacheribbe, Andrei Cristea, Eljas Roellin, Dominique Von Schiller, Eve Coomber, May Hu, Gabriella Ivanova, Fereshteh Torabi, Nima Subasinghe, Laura Durrant, Vahid Shariati, Chiara Chiereghin, Michela Colombo, Ilaria Gori, Federica Santonastaso, Matilde I. Conte, Cecilia Dominguez Conde, Alice Giustacchini, Genes & Health Research Team, Nicole Staudt, John Wright, Richard Trembath, Craig A. Glastonbury, Julien Gagneur, Sarah Finer, Brian Clarke, Marc Jan Bonder, Fabian J. Theis, Fabiola Curion, Vivek Iyer, Oliver Stegle, Francesco Paolo Casale, Sarah A. Teichmann, David A. van Heel, Nicola Pirastu, Blagoje Soskic, Nicole Soranzo

## Abstract

Human immune composition reflects genetic background and cumulative life-course exposures, yet how these influences shape immune architecture across populations and disease remains poorly understood. Here we integrate single-cell transcriptomics, matched clinical haematology and electronic health records from 5,079 individuals of European, British-Pakistani and British-Bangladeshi ancestries to quantify 26 circulating immune cell types and 62 transcriptional states. We map immune compositional variation associated with age, sex, adiposity and smoking, revealing distinct patterns across adulthood and associations between modifiable exposures and advanced immune age. Across common diseases, immune compositional signatures converge on recurrent inflammatory myeloid, adaptive-memory and interferon programmes, whose expression varies with age and population context. Cardiovascular therapies are associated with selective shifts in disease-associated immune states. Immune composition further discriminates common cardiometabolic diseases and resolves patients into endotypes with distinct metabolic and clinical profiles. These findings establish circulating immune architecture as an integrative phenotype linking life-course exposures, population context and disease, providing a framework for precision population immunology.

## Introduction

The immune system is central to human health, defending against pathogens, surveilling for malignant transformation, and regulating inflammation while maintaining tolerance to self. Beyond inherited factors^1–3^, circulating immune-cell composition is shaped by cumulative life-course exposures and demographic factors such as age, sex and ancestry, and environmental influences such as adiposity, smoking and reproductive status^4–7^. Most work dissecting this variation has isolated single factors, such as age, in controlled settings. Such studies cannot capture the joint effects of genetic background, life-course exposures and population context, so the combined contribution of these factors to immune variation remains poorly characterised, particularly across diverse populations.

Complete blood counts (CBCs) transformed clinical medicine by enabling quantitative assessment of blood composition, flagging deviations from reference ranges that signal infection, inflammation or haematological disease. Yet blood cell composition varies substantially among healthy individuals and populations for reasons that remain incompletely understood^4,8,9^, and conventional haematology collapses circulating immune cells into a handful of broad lineages, limiting resolution of finer scale phenotypes. Flow cytometry improves resolution, but predefined marker panels and differences in experimental protocols limit the breadth and comparability of immune phenotyping across cohorts^10,11^.

Single-cell transcriptomics overcomes these limitations, enabling unbiased, high-resolution characterisation of immune cell types and states across large sample sizes^12–16^ and capturing cellular diversity invisible to CBCs and impractical to scale with FACS. Because single-cell genomics yields relative rather than absolute cell proportions, comparisons across individuals are inherently compositional: a change in one population necessarily shifts the relative abundance of others. Applied at population scale, however, single-cell technologies provide a detailed view of immune architecture and support rigorous modelling of how biological and environmental factors jointly shape immune composition across the life course.

Pairing single-cell genomics with large, population-representative cohorts and linked electronic health records (EHRs) offers a new way to model the relative contribution of demographic, epidemiological and disease-related determinants of immune variation across life^17,18^. By enabling hypothesis-free investigation across multiple disease diagnoses, such resources can reveal how common diseases reshape immune-cell composition, uncover underappreciated immune mechanisms contributing to the pathogenesis of classically non-immune diseases, and identify immunologically distinct patient subgroups with potential relevance for disease stratification. They further provide opportunities to systematically evaluate emerging risk factors and identify early biomarkers of disease onset, particularly where multiple chronic conditions coexist.

Here, we leverage a single-cell molecular phenotyping resource of the immune system spanning thousands of participants of European, British-Pakistani and British-Bangladeshi ancestry (Giacopuzzi et al. 2026) to characterise population-level variation in peripheral blood mononuclear cell (PBMC) types at high resolution, moving beyond canonical CBC traits to finely resolved immune phenotypes. By integrating single-cell profiles with matched haematology and EHR data, we define peripheral immune composition across the adult life course, identify its population- and age-related drivers, and reveal disease-associated immune states with relevance to drug targeting, disease discrimination and endotyping. Together, these analyses establish single-cell genomics as a scalable platform for population immunology and precision medicine.

## Results

### High-resolution scCBCs enable population-scale quantification of immune-cell cross-sectional age profiles

We analysed CARDINAL, a single-cell genomics resource comprising ∼6,000 individuals recruited through two UK population cohorts: 4,754 European-ancestry participants from UK Biobank (UKB) and 1,248 British-Pakistani and British-Bangladeshi participants from Genes & Health (G& H) (Giacopuzzi et al.). The two cohorts analysed here exhibit distinct age distributions (UKB: 53–85 years; G& H: 16–89 years; **Supplementary Table 1**), enabling complementary resolution of age-associated changes in immune architecture across the life course. The dataset comprises approximately 9.8 million peripheral blood mononuclear cells (PBMCs) collected from fresh blood samples, together with matched Sysmex haematology measurements (**Methods**). Participants contributed a median of 1,471 (interquartile range 1,069 - 1,923) PBMCs per donor in UKB and 1,738 (interquartile range 1,268 - 2,367) in G& H.

To derive absolute immune-cell counts, we developed single-cell CBCs (scCBCs) by scaling single-cell subtype proportions to donor-matched Sysmex lymphocyte and monocyte counts (**Methods; Supplementary Fig. 1**). This yielded estimated absolute counts for 26 immune cell *types* (cell type 2, CT2) and 62 cell *states* (CT3). After restricting analyses to samples with concordant lymphocyte-to-monocyte ratios across the two platforms (**Methods; Supplementary Fig. 2**), the final dataset comprised 4,094 UKB and 985 G& H participants, corresponding to 86% and 79% of the initially profiled participants respectively (**Supplementary Table 1, Fig. 1a**). The concordance criterion was defined as exclusion of samples with a Sysmex-to-single-cell lymphocyte-to-monocyte ratio lying outside [Q_1_ − 5*IQR, Q_3_ + 5*IQR]; the two platforms showed proportional scaling (R=0.86 for UKB and R=0.84 for G& H; **Methods; Supplementary Fig. 2**). By anchoring single-cell proportions to clinical blood counts, scCBCs enable quantitative comparisons of immune-cell abundance estimates across individuals and mitigate compositional bias inherent in single-cell data^19^. Consistent with this, proportion-based analyses showed the expected negative correlation bias, whereas this artefactual dependency was not observed in scCBCs (**Supplementary Fig. 3–4**).

**Figure 1:**
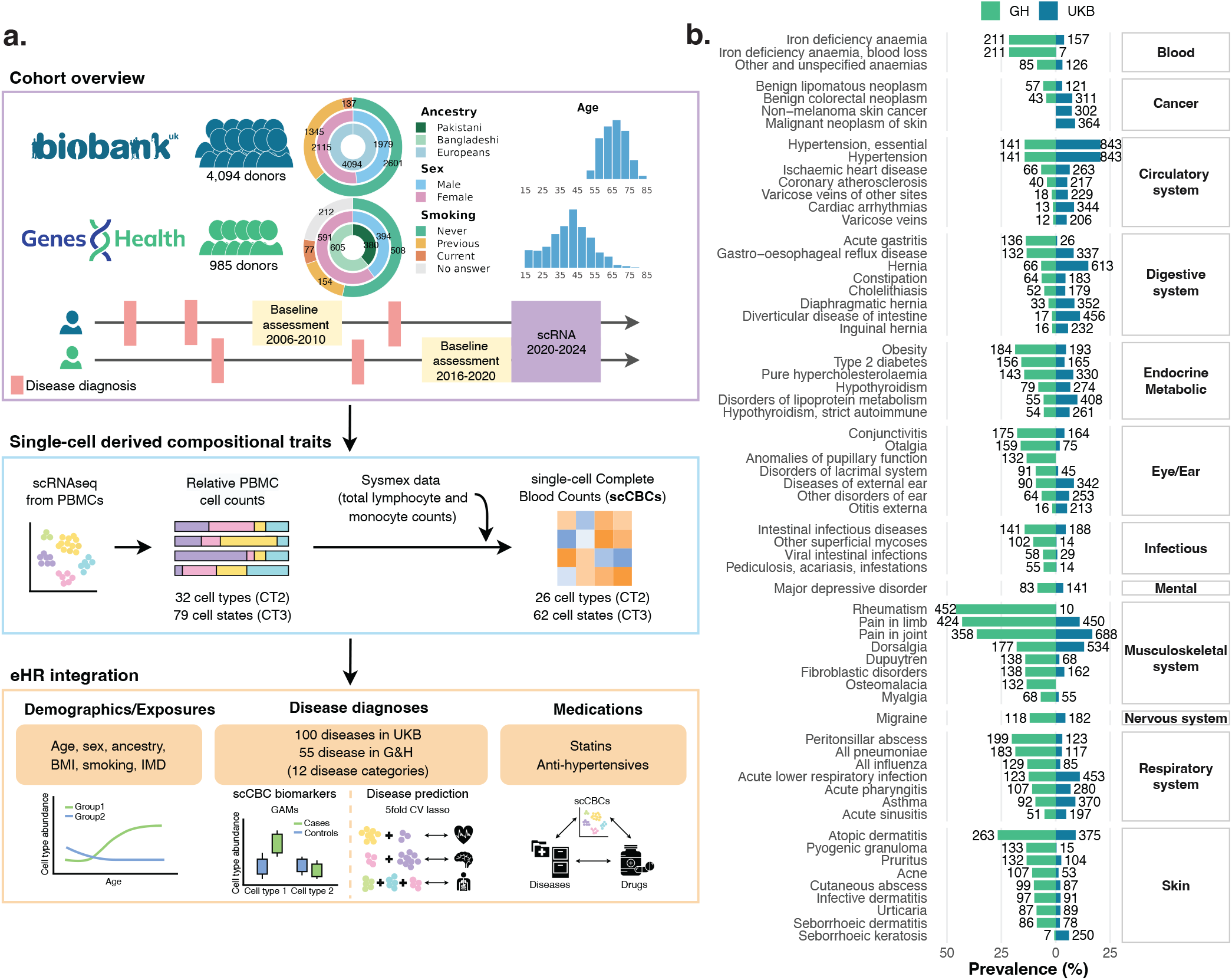
Study design and disease prevalence across cohorts. **a,** Overview of the study design. The top panel summarises the two contributing cohorts, UK Biobank and Genes & Health, including donor numbers, demographic characteristics and timing of baseline assessments and peripheral blood mononuclear cell (PBMC) single-cell RNA sequencing (scRNA-seq). The middle panel shows derivation of relative PBMC cell counts from scRNA-seq data and their integration with Sysmex total lymphocyte and monocyte counts to generate single-cell complete blood counts (scCBCs). The bottom panel outlines downstream analyses of cell-type abundance and scCBC traits in relation to age, diseases and medications. **b,** Disease prevalence in Genes & Health (left) and UK Biobank (right). Each row denotes a disease with prevalence of at least 5% in at least one cohort.

We next used scCBCs to quantify the associations between demographic and lifestyle factors on immune composition by modelling changes in scCBCs jointly as a function of age, sex, population background, body mass index (BMI) and smoking, hence estimating the mutually adjusted contribution of each variable (**Methods**). Due to personal data privacy restrictions (including UK-GDPR), the biomedical variables of the two cohorts are held in separate Trusted Research Environments, and therefore all analyses integrating single-cell data with clinical phenotypes were performed independently within each cohort. Nevertheless, this approach allowed us to quantify variable-associated changes in each cell state and compare their magnitude and direction across cohorts.

Associations with age recapitulated in both cohorts the profound remodelling of the immune compartment (**Fig. 2a, Supplementary Tables 3-4**), with over half of the 62 cell states (CT3 level) showing significant association with age (32/62 in UKB and 34/62 G& H; FDR < 0.05). These included progressive depletion of naive B cells^20,21,22^, naive CD4^+^ and naive CD8^+^ T cells^23^, mucosal-associated invariant T (MAIT) cells, γδ T cells and ILC3s^21^, alongside expansion of CD14^+^ and CD16^+^ monocytes, adaptive CD16^+^ NK cells, and increased CD4^+^ Treg effector populations and terminally differentiated EOMES^+^, NCAM1^+^ and ZNF683^+^ CD8 TEMRA states^23^. For each cell state an age-related effect was defined from standard deviations in scCBC change per 10-year increase in age. Despite the age difference for the two cohorts, the magnitude of age-associated changes was broadly comparable (median −0.06, range −0.48 to 0.20 in UKB and median −0.05, range −0.45 to 0.23 in G& H), with 18 of 62 cell types showing FDR-significant, directionally concordant associations. Restricting G& H to the shared age range (53+ years), 7 of its 10 significant associations were directionally concordant in UKB (**Supplementary Table 5**). Naïve early CD8^+^ T cells showing the greatest age-associated decline in both cohorts (β = −0.48 in UKB and β = −0.45 in G& H; 95% CI [−0.50, −0.46] and [−0.48, −0.42]; FDR-adjusted p = 5.75 x 10^−105^ and 2.8 x 10^−44^, respectively). The broader age distribution of the G& H cohort (16–89 years) provided a finer assessment of stages of compositional variation during the pre-immunosenescence stage^24^, highlighting more dynamic patterns of variation in subgroups of cells (**Fig. 2a, Supplementary Fig. 5-6, Supplementary Tables 3-4**). Naive lymphocytes, MAIT cells and several innate lymphoid populations showed monotonically declining counts throughout adulthood, while inflammatory monocytes and differentiated CD4^+^ and CD8^+^ T-cell states showed corresponding monotonically increasing variation. Further, non-linear population variation was observed for other subsets (**Fig. 2b; Supplementary Fig. 5-6**), including for instance CD4^+^ central-memory T cells that increased until midlife and declined thereafter, with an estimated inflection point at approximately 52 years (FDR for non-linearity = 0.012)^21^. The extended age range also enabled robust quantification of rare populations, revealing age-associated depletion of ILC2s, innate lymphoid progenitors and plasmablasts, populations that conventional haematology cannot reliably resolve (**Fig. 2a**). Therefore, by averaging effects across individuals, these population-level patterns inferred from cross-sectional data inform understanding and quantitative assessments of age-associated longitudinal trajectories, complementing information from cell atlasing studies^20,21^.

**Figure 2:**
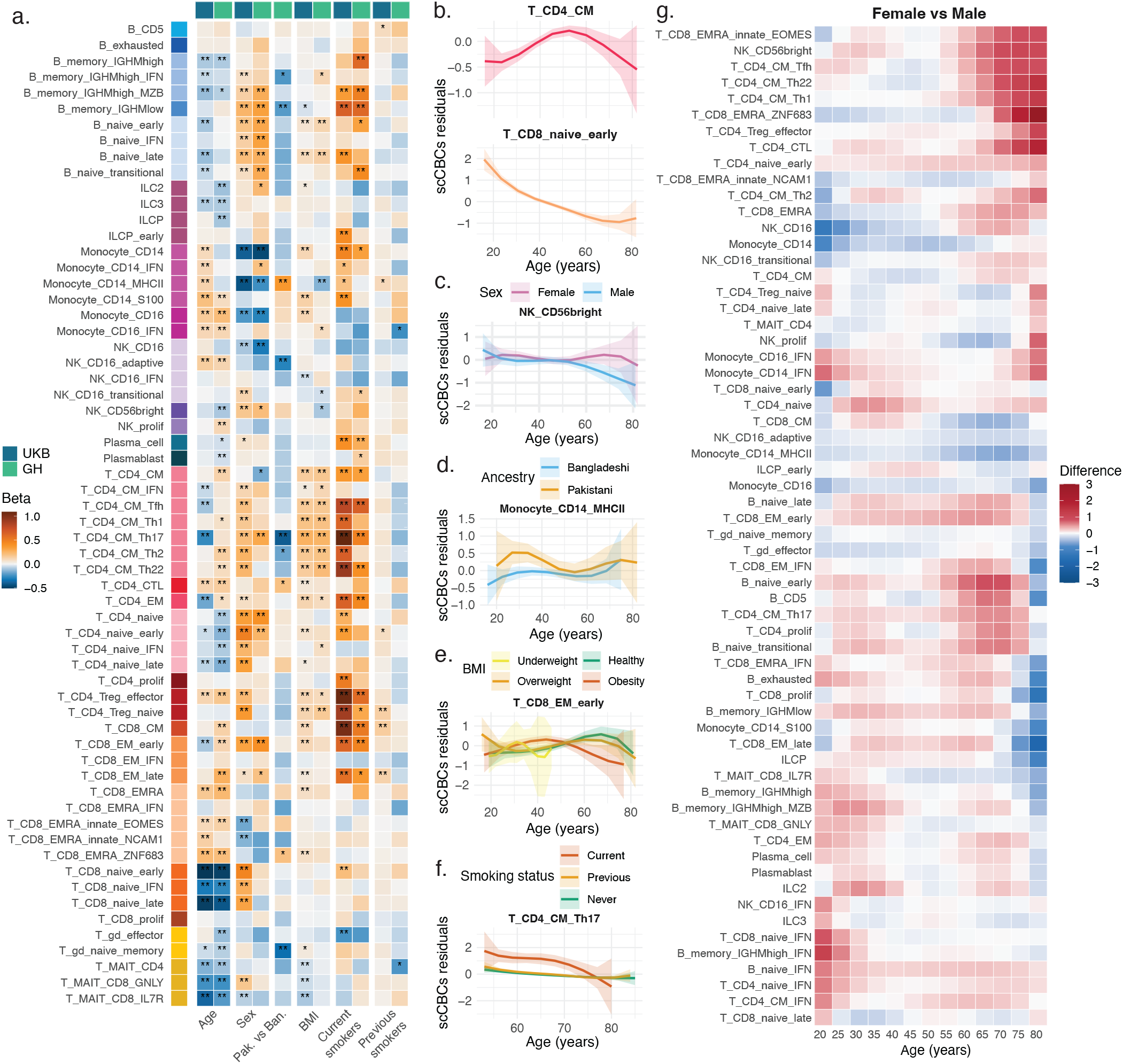
Associations and age-associated immune cell profiles in UK Biobank and Genes & Health. **a,** Heatmap of associations between immune cell abundance at CT3 level and sex, age, ancestry, body mass index (BMI) and smoking status in UK Biobank (UKB; n=4,083) and Genes & Health (G& H; n=710). Colours indicate beta coefficients from linear models, representing the expected change in standardised cell abundance associated with each predictor; positive values indicate higher abundance and negative values indicate lower abundance. A single asterisk denotes nominal significance (*p <* 0.05) and double asterisks denote significance after Benjamini–Hochberg correction. **b–f,** Examples of smoothed age profiles of CT3 scCBC residuals, stratified by sex (**c**), ancestry (**d**), BMI (**e**) and smoking status (**f** ). scCBC residuals were derived by correcting scCBCs for all covariates not used for stratification in each panel. Shaded bands indicate 95% confidence intervals. **g,** Heatmap of age-specific sex differences in risk-factor-corrected immune cell-type abundances in G& H. Each tile shows the contrast between the smoothed age profiles of females and males at each age for each cell type. Smoothed age profiles were derived from cell abundances corrected for covariates not used for stratification. Cell types are ordered by hierarchical clustering of their standardised age-specific contrast profiles, based on Pearson correlation distance and average linkage. Colours represent the magnitude and direction of the female-versus-male difference, with positive values indicating higher abundance in females and negative values indicating higher abundance in males.

We recapitulated the established immune dimorphism between sexes (**Fig. 2a, Supplementary Tables 3-4**) with greater abundance of adaptive immune populations in females — naive and memory B cells, naive CD4^+^ T cells and several CD8^+^ memory subsets^25,26^ — while males showed increased CD14^+^, CD16^+^ and MHCII^high^ monocytes together with cytotoxic CD16^+^ NK cells^27^. The resolution of our atlas further enabled identification of previously unreported associations, including increased abundance of early effector memory CD8^+^ T cells and depletion of IL7R^+^ MAIT CD8^+^ cells in UKB females, suggesting selective sensitivity of these subsets to cohort structure or annotation depth. For cell states with FDR < 0.05, the magnitude of sex difference was greater in G& H (median = 0.26 s.d., range −0.43 to 0.37) than UKB (median = 0.17 s.d., range −0.40 to 0.44), possibly due to differences in age structure between the cohorts. CD14^+^ monocytes showed the greatest difference between sexes, with counts 0.37 s.d. and 0.43 s.d. higher in males than in females in UKB and G& H respectively (FDR-adjusted p = 2.31 x 10^−32^ and p = 3.76 x 10^−5^). These sex differences were highly variable across adulthood at the population level^28^. Across the 62 immune states, we observed persistent differences, progressive divergence and attenuation of sex effects when averaging between donors of similar age (**Supplementary Fig. 7-8**). Innate populations displayed particularly pronounced age-associated patterns in G& H (**Fig. 2g**). CD16^+^ NK cells showed a significant overall age-dependent sex difference (FDR-adjusted p = 1.59×10^−3^), with greater counts in males supported until approximately age 30. Early naive B cells also showed significantly different sex-specific age patterns (FDR-adjusted p = 0.025), with higher abundance in females supported from approximately age 57 onward. CD56^bright^ NK cells showed suggestive evidence of differing age patterns by sex (p = 0.010), with higher abundance in females supported from approximately age 60 onward (**Fig. 2c, Supplementary Fig. 7–8**). These age-associated patterns help explain the cohort differences: UKB’s older age distribution predominantly sampled ranges where sex differences were already established, while G& H’s broader age range captured their emergence across adulthood. Together, these analyses show that age- and sex-related associations are heterogeneous and cell-state specific, with sex differences that can strengthen, weaken, or reverse over the adult life course.

Extending this framework to fine-scale population variation, scCBCs uncovered previously uncharacterised structure within the G& H cohort. British-Pakistani and British-Bangladeshi donors represent two South Asian groups with distinct genetic and environmental backgrounds, including differences in autozygosity, population structure and consanguinity-associated founder effects^29^. After adjustment for the other covariates, CD14^+^ MHCII^high^ monocytes were enriched in British-Pakistani donors, whereas IGHMlow memory B cells, adaptive CD16^+^ NK cells, CD4^+^ central-memory Th17 cells and naive-memory γδ T-cell subsets were enriched in British-Bangladeshi, indicating coordinated redistribution across adaptive and barrier-associated compartments (**Fig. 2a, Supplementary Tables 3-4**). These differences persisted variably across donor ages: while CD4^+^ central-memory Th17 and naive-memory γδ T cells showed significantly different age-related patterns between ancestry groups across all ages (FDR-adjusted p = 6.19×10^−3^ for both), CD14^+^ MHCII^high^ monocyte diversification was restricted only up to approximately age 41 (FDR-adjusted p = 0.022; **Fig. 2d, Supplementary Fig. 9**). This pattern may reflect greater sensitivity of the myeloid compartment to early-life infectious exposures, with more persistent effects in adaptive and barrier-associated compartments^30,31^.

We finally investigated socioeconomic status (SES) as proxied by the index of multiple deprivation (IMD), a composite measure of area-level deprivation capturing income, education, employment, health, housing, living environment, crime and access to services. IMD showed no robust associations with scCBCs in either cohort (FDR < 0.05; **Supplementary Fig. 10a-b**). Nominal associations (p < 0.05) nevertheless suggested subtle, cohort-specific effects on lymphoid and cytotoxic homeostasis. In UKB, IMD was positively associated with BMI and smoking (**Supplementary Fig. 10c–f**), indicating that SES gradients may act largely through modifiable exposures such as adiposity and smoking. The recapitulation of established patterns of immune ageing and sexual dimorphism supports the reliability of scCBCs for quantifying immune-cell abundance at population scale, while providing substantially greater resolution than conventional haematology. By distinguishing 26 cell types and 62 functional cell states, scCBCs reveal how demographic factors shape immune composition across diverse population contexts.

### Smoking and BMI remodel immune-cell composition across adulthood

Age and sex establish baseline immune composition through developmental and hormonal signalling, but lifestyle factors and exposures further shape immune homeostasis throughout life^32,33^. Smoking and adiposity are major determinants of chronic inflammation and have both been implicated in accelerated immune ageing^7,34–36^. We therefore asked how these two common exposures were associated with immune-cell composition in models accounting for age, sex and population background (**Methods**). To account for incomplete data at the time of PBMC collection, the most recent available BMI and smoking measures were taken as proxies, after confirming their temporal stability between baseline and recall measurements (**Methods; Supplementary Fig. 11**).

Smoking was primarily associated with changes in the adaptive immune compartment (**Fig. 2a, 3b, Supplementary Tables 3-4, Supplementary Fig. 12-13**). Current smokers showed elevated counts of memory B cells, central-memory CD4^+^ subsets, memory CD8^+^ cells, regulatory T cells^26,37^, together with naïve B cells and plasma cells, associations that have not been previously reported to our knowledge. In UKB, smoking was further associated with naïve T cells, classical and S100^+^ CD14^+^ monocytes, early ILCPs and depletion of γδ effector T cells^38^, as well as several corresponding cell states. We also identified previously unreported expansion of transitional/late naïve B cells, plasma cells, consistent with tobacco-driven activation of the naïve B-cell pool and differentiation towards antigen-experienced states^32^. Immune cell composition in former smokers largely converged with that of never-smokers, with modest residual differences confined to regulatory and memory T-cell populations (**Fig. 2a, 3b, Supplementary Tables 3-4**), potentially indicative of recovery of immune cell composition following smoking cessation^37^. Our quantitative assessment in this population setting implicated 24 affected cell types in individuals classified as current versus never smokers in UKB, and 12 in G& H (FDR < 0.05), with median effects of 0.49 s.d. (range −0.22 to 0.99) in UKB and 0.44 s.d. (range 0.25 to 0.59) in G& H, compared with only up to three cell types differentiating former from never smokers in UKB, with small effects (range 0.11 to 0.13 s.d.).

Adiposity (as proxied by BMI in this study) showed broader and more heterogeneous effects, implicating 25 of 62 cell states in UKB and 10 of 62 cell states in G& H (FDR < 0.05; **Fig. 2a, Supplementary Tables 3-4**). Median scCBCs changes were smaller in UKB (0.09 s.d., range −0.09 to

0.23 per one–standard-deviation increase in BMI) than in G& H (0.16 s.d., range −0.14 to 0.23). At finer resolution, we observed expansion of early and late naïve B cells, CD4 central memory subsets spanning Tfh, Th1, Th2, Th17 and Th22 lineages, and naïve Tregs (**Fig. 2a, Supplementary Tables 3-4**), extending prior work on obesity-linked broad B-cell and T-cell remodelling by resolving the specific cell states involved^34^. Further, NK CD16+ IFN+ and GNLY+/IL7R+ MAIT subsets all decreased with BMI in UKB, providing finer cell-state resolution relative to earlier reports of obesity-associated changes in NK and MAIT compartments^39^ and suggesting a more selective erosion of innate-like immune surveillance in older individuals^40^. BMI-associated effects were strongly correlated with donor age: individuals classified as underweight, overweight or with obesity followed distinct distributions with age, with divergence from healthy-weight individuals becoming more pronounced at older age (**Fig. 3a, Supplementary Fig. 14-15**). Sex-stratified analyses revealed broadly similar BMI-associated patterns in females and males (**Supplementary Fig. 16**). CD14^+^ MHCII^high^ monocyte counts were higher in healthy-weight compared to overweight and obese donors from approximately age 53 onward (Holm-adjusted p = 1.6×10^−3^ and 1.2×10^−3^, respectively; BMI FDR-adjusted p = 0.0123). CD4^+^ central-memory Th1, Th17 and Th22 cells also showed significant BMI-dependent age patterns (FDR-adjusted p = 0.0123 for each), with higher abundance in obese individuals during early-to-mid adulthood relative to both healthy-weight and overweight groups. Early effector-memory CD8^+^ T cells showed a similar but weaker pattern (nominal p = 8.0×10^−3^), with higher abundance in obese individuals supported between approximately age 33 and 46 (**Fig. 2e**). These population patterns support widespread age-dependent remodelling of immune compositional architecture with adiposity^34^.

**Figure 3:**
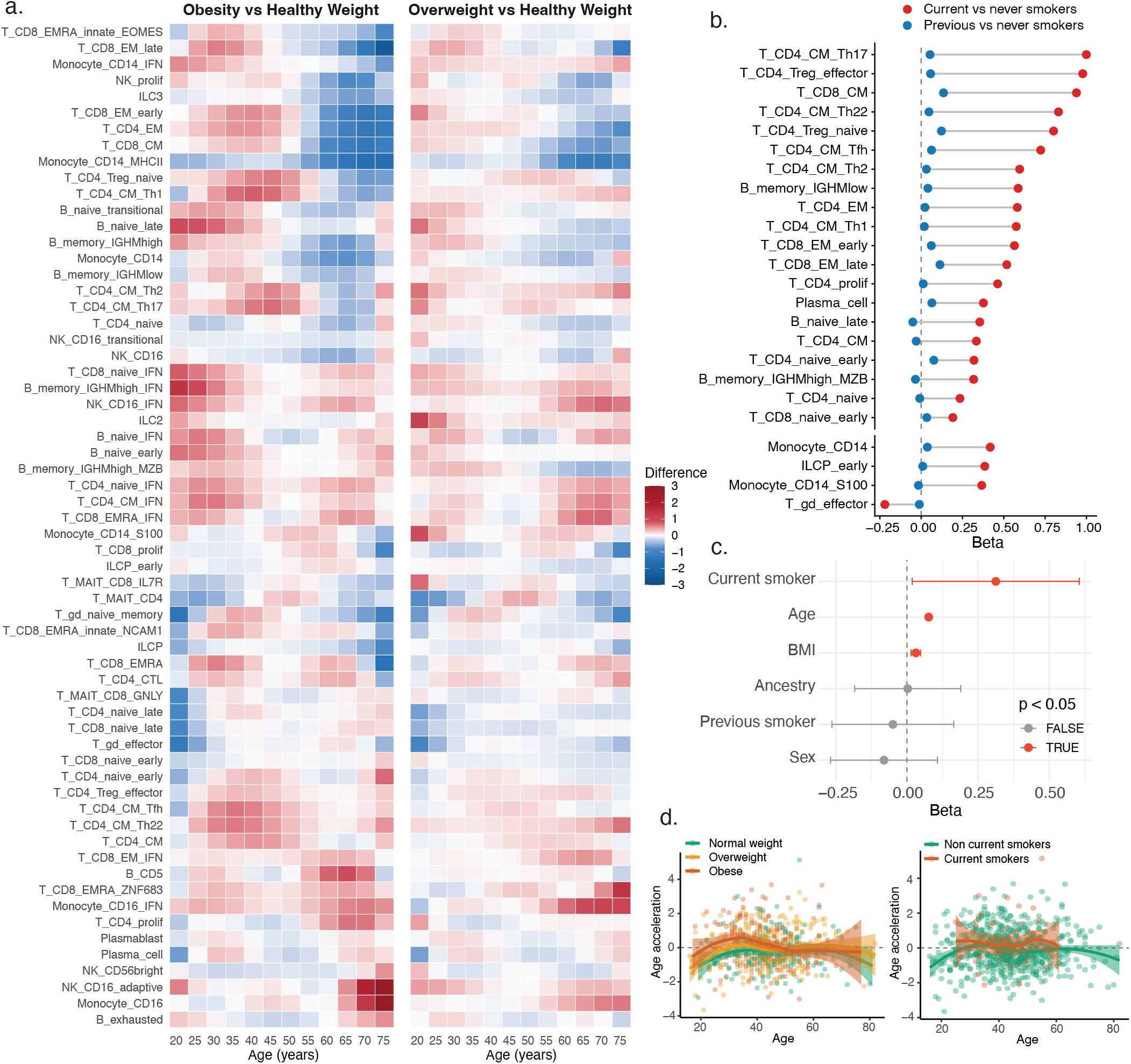
BMI and smoking shape immune cell-state composition and cellular ageing. **a,** Heatmap of age-specific differences in risk-factor-corrected immune cell-type abundances according to BMI in Genes & Health. Each tile shows the contrast between the smoothed age profiles of individuals with obesity or overweight and those with healthy weight at each age for each cell type. Smoothed age profiles were derived from cell abundances corrected for covariates not used for stratification. Cell types are ordered by hierarchical clustering of their standardised age-specific obesity contrast profiles, based on Pearson correlation distance and average linkage. Colours indicate the magnitude and direction of the difference, with positive values denoting higher abundance in obesity/overweight and negative values denoting lower abundance. Overweight profiles showed the same overall pattern as obesity, but with smaller effect sizes. **b,** Dumbbell plot of smoking-associated changes in immune cell-type abundances in UKB. Beta coefficients from associations between CT3 scCBCs and risk factors are shown for current versus never smokers (red) and previous versus never smokers (blue). Cell types are grouped as adaptive (top) or innate/innate-like (bottom). Current smoking was associated with stronger effects in adaptive cell types, whereas associations in innate/innate-like cell types were more restricted; previous smoking showed attenuated effects. **c,** Forest plot of covariate associations with PLS1, the primary cellular ageing axis derived from CT2 scCBCs in Genes & Health. Positive coefficients indicate a shift towards an immune profile characteristic of older individuals. PLS1 was associated with age, BMI and current smoking status. **d,** Age-adjusted PLS1 residuals, defined as the residual from regressing PLS1 on chronological age, plotted across BMI categories (left) and smoking status (right). Positive values indicate an older-than-expected age-associated immune profile relative to individuals of the same chronological age. Curves were fitted using loess and are shown with 95% confidence intervals.

We next sought to unpick the observed correlation between immune cell populations, smoking, BMI and age. Of 32 (UKB) and 34 (G& H) age-associated cell states, 16 and 3 were also associated with BMI and 10 and 5 with current vs never smoking (FDR < 0.05). To quantify immune ageing, we derived an immune age-score in G& H (owing to its wider age distribution) using partial least squares (PLS) regression on scCBCs (**Methods**). The first latent component (PLS1) captured the major age-associated axis of compositional variation, explaining 41.5% of chronological age variance (**Supplementary Fig. 17b**). Older immune profiles were characterised by depletion of naive lymphocyte and innate-like populations together with expansion of memory, terminal-effector and inflammatory myeloid states (**Supplementary Fig. 17c**). After adjustment for chronological age and demographic variables, both current smoking (p = 3.70 x 10^−2^) and higher BMI (p = 2.54 x 10^−4^) were independently associated with increased age deviation, whereas former smoking showed no significant association (**Fig. 3c-d**).

Our population data therefore captured distinct modes of immune remodelling mediated by modifiable risk factors, confirming the deep perturbation of adaptive immune populations associated with smoking, and the broader and age-dependent involvement of adaptive and innate compartments with BMI. Despite these differences, both current smoking and higher BMI were associated with an older age-associated immune profile, highlighting immune composition as a potential quantitative marker of exposure-associated immune variation.

### Shared immune programmes connect disease, population context and life-course immune remodelling

Immune composition integrates the cumulative effect of genetic background, modifiable exposures and ageing, providing a systems-level readout of host physiology. We therefore asked whether scCBCs could identify recurrent immune programmes associated with prevalent disease across two population cohorts with distinct demographic composition and disease burden. To characterise the disease landscape of the two cohorts, we first quantified phecode-defined conditions with ≥5% prevalence in either cohort, encompassing twelve major disease categories (**Fig. 1b, Supplementary Table 2**). UKB participants exhibited lower prevalence of metabolic disorders and smoking, whereas the G& H cohort showed greater burden of infectious, respiratory, dermatological and metabolic diseases (**Supplementary Table 2**). Conversely, cardiovascular and neoplastic diseases were more frequent in UKB. These differences recapitulated established epidemiological patterns^41^ and emphasised the need to account for population context when interpreting immune variation (**Supplementary Fig. 18**).

We next modelled immune cell abundance for 100 and 55 disease diagnoses in UKB and G& H (≥35 cases/disease; **Supplementary Table 2**) using generalised additive models adjusted for age, sex, ancestry, BMI and smoking (**Methods**).

These signatures were summarised as a disease network, with edges capturing similarity between disease–cohort pairs based on their associated immune-cell states (≥2 shared states with concordant directions; Jaccard ≥0.2; **Methods**). Disease-associated immune signatures organised into coherent regions of the immune space, with related conditions clustered together, despite their clinical diversity (**Fig. 4a**). Rather than generating disease-specific immune architectures, many conditions converged on a limited number of recurrent cell-state programmes, indicating that diverse diseases remodel common components of the circulating immune system.

**Figure 4:**
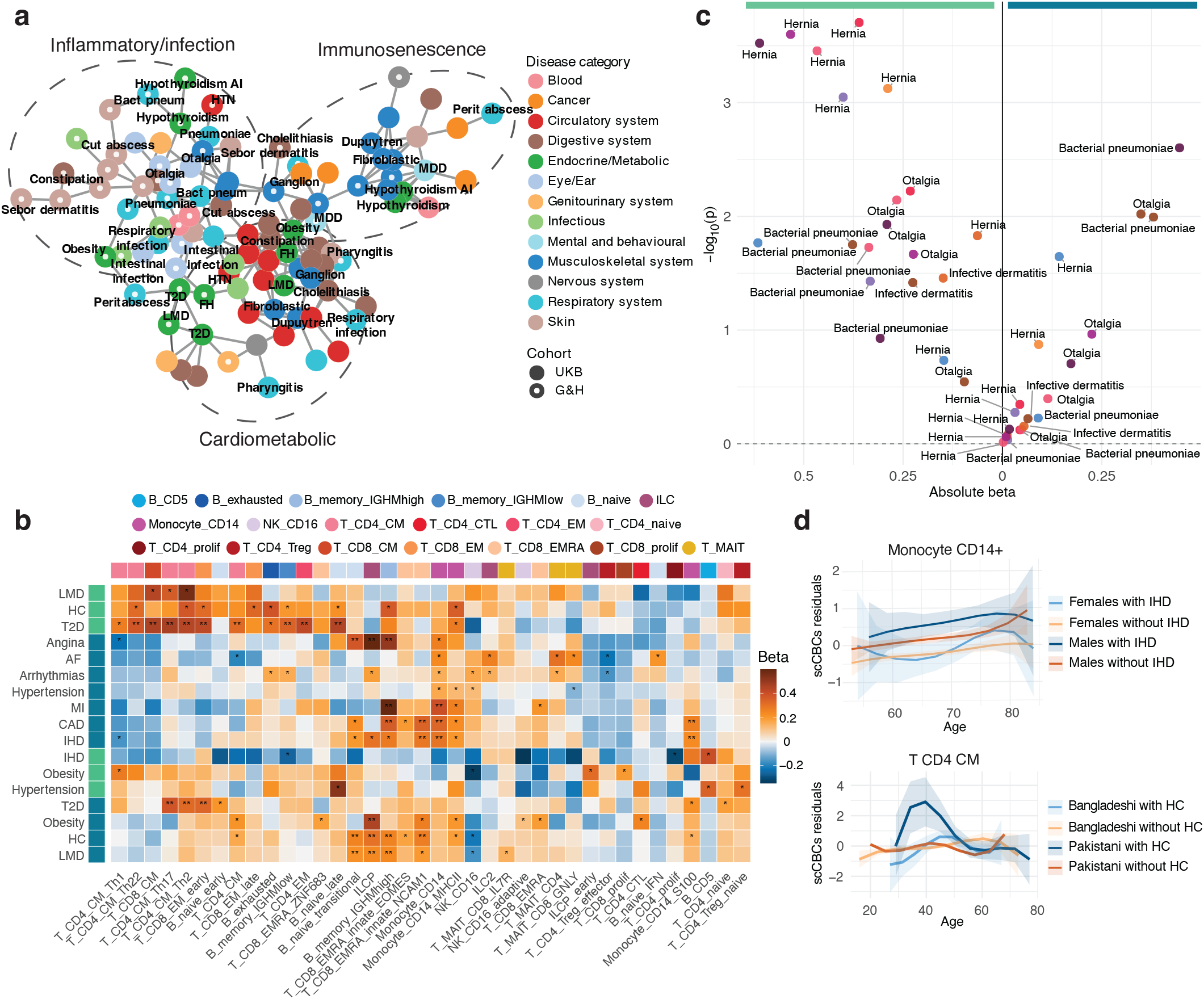
Disease-associated cell-state programmes across cohorts. **a,** Disease–cell signature network based on Jaccard similarity of signed, significant cell-type level 3 states across UK Biobank (UKB) and Genes & Health (G& H). Each node represents a disease–cohort pair; and edges connect disease pairs sharing signed significant cell-type signatures (Jaccard similarity *>* 0.2, with at least two shared cell types). Disease labels are shown only for diseases present in both cohorts, to indicate whether their corresponding nodes cluster near one another. **b,** Heatmaps of regression coefficients (*β*) from generalised additive models relating CT3 scCBCs to disease status for cardiometabolic traits. Colour encodes the magnitude and direction of the *β* coefficient, with blue indicating lower abundance in cases and orange indicating higher abundance in cases. Asterisks denote statistical significance: ∗ *p <* 0.05; ∗∗ Benjamini–Hochberg false discovery rate *<* 0.05. **c,** Volcano plot showing associations between CT3 scCBC and multiple diseases (bacterial pneumoniae, hernia, otalgia, infective dermatitis) in Genes & Health (left) and UK Biobank (right), as example of G& H interferon-response signatures. Each point represents an IFN scCBC trait; the x axis shows the absolute beta coefficient and the y axis shows −*log*10(*p*). The highlighted signatures are the significant ones (*p <* 0.05). Point colours represent cell types, with the corresponding colour legend shown in panel b. Only traits significant in at least one cohort (*p <* 0.05) were retained. **d,** Smoothed age profiles of scCBC residuals across age for representative cell types and disease contrasts, fitted using B-splines and shown separately by disease status. Top panel shows CD14+ monocyte abundance in UKB, stratified by ischaemic heart disease and sex, showing male-specific age-dependent enrichment in affected individuals. Bottom panel shows CD4 central memory T-cell abundance in G& H, stratified by hypercholesterolaemia and ancestry, with higher abundance at younger ages in British-Pakistani participants with disease and smaller differences between groups after mid-life. Age profiles were adjusted for covariates not used for stratification, including sex, BMI, smoking and ancestry as appropriate; shaded bands denote 95% confidence intervals.

The most prominent programme linked several cardiometabolic diseases in UKB, and was dominated by inflammatory myeloid remodelling (**Fig. 4b; Supplementary Fig. 19–22, Supplementary Tables 6-7**). Across coronary atherosclerosis, ischaemic heart disease, hypertension and related disorders, we observed coordinated expansion of CD14^+^ inflammatory monocytes, including S100^+^ and MHCII^high^ states, together with innate lymphoid progenitors and terminally differentiated CD8 EMRA populations, with the most extreme association implicating ILCPs in angina pectoris (β = 0.59 s.d. higher ILCP counts in individuals with disease compared to unaffected individuals; FDR-adjusted p = 1.27×10^−3^). These states are consistent with established roles in atherogenesis and vascular inflammation^42,43^ and with cytotoxic T-cell involvement in disease progression^44^. Metabolic diseases showed contrasting immune signatures across cohorts (**Fig. 4b**). In UKB, lipoprotein disorders shared lymphoid features of cardiovascular disease but lacked inflammatory CD14^+^ monocyte signals; pure hypercholesterolaemia additionally enriched innate-like CD8^+^ EMRA states. Obesity combined monocyte and cytotoxic lymphoid changes with pronounced CD4^+^ central memory and adaptive NK-cell expansion, without B-cell remodelling. Type 2 diabetes predominantly affected CD4^+^ and CD8^+^ populations, alongside increased S100^+^ CD14^+^ monocytes^45,46^. In G& H, hypercholesterolaemia and type 2 diabetes shared prominent B-cell remodelling, including late naïve and IGHMlow memory states, together with MHCII^high^ CD14^+^ monocytes and memory T-cell expansion, with diabetes showing broader T-cell changes^47^. Lipoprotein disorders primarily involved memory T-cell subsets. G& H obesity was distinguished by increased early ILCP, Th1-like CD4^+^ central memory and proliferating CD8^+^ cells.

A second programme captured shared features of immunosenescence across diverse conditions. In UKB, hypothyroidism, major depressive disorder and psoriasis were united by depletion of early and late naïve CD4^+^ and CD8^+^ T-cell subsets, with disease-specific remodelling of memory, humoral and myeloid states: MAIT CD8^+^ GNLY^+^ cells were reduced in hypothyroidism; IGHMhigh memory B cells and CD14^+^ S100^+^ monocytes were expanded in major depressive disorder; and plasmablasts were increased in psoriasis (**Supplementary Fig. 19-20, Supplementary Tables 6-7**). In G& H, musculoskeletal and pain syndromes (dorsalgia, fibroblastic disorders, limb or joint pain, Dupuytren’s disease, rheumatism) showed a related signature, marked by loss of late naïve CD4^+^ and CD8^+^ T cells, reduced CD4^+^ central-memory Th17 and effector-memory populations, and depletion of early naïve B cells, alongside increased NK CD16^+^ adaptive and transitional subsets in limb pain and rheumatism (**Supplementary Fig. 21–22, Supplementary Tables 6-7**). This axis points to contraction of naïve lymphocyte pools with compensatory shifts in memory and innate compartments, in line with established hallmarks of immunosenescence^24,48^.

Third, many infectious and barrier-associated phenotypes in G& H converged on a pervasive type I interferon (IFN) programme spanning myeloid, NK, T- and B-cell compartments (**Fig. 4c, Supplementary Fig. 21–22, Supplementary Tables 6-7**). IFN-activated states were shared across respiratory, gastrointestinal, cutaneous and musculoskeletal conditions, suggesting that recent or persistent immune activation induces common interferon-responsive cell states irrespective of the affected tissue^49,50^. These findings indicate that clinically distinct diseases frequently converge on a small number of conserved immune programmes reflecting inflammatory, adaptive-memory and antiviral responses.

Finally, we asked whether common diseases were associated with distinct patterns of immune-cell abundance across age groups at the population level. We modelled cross-sectional age profiles using B-splines, stratifying by disease status, sex, BMI and population background (**Methods**). Across multiple conditions, disease-associated differences varied with age and depended on sex, adiposity and ancestry, suggesting changes in the timing and magnitude of immune remodelling rather than uniform shifts in cell abundance.

Two representative examples illustrate these disease-associated differences in age profiles. In UKB, the elevation in CD14^+^ monocyte frequencies among males with ischaemic heart disease became more pronounced at older ages, whereas affected females showed age-associated patterns similar to unaffected controls, suggesting sex-specific remodelling of the innate compartment in heart disease ^51,52^ (**Fig. 4d**). In G& H, hypercholesterolaemia was associated with higher CD4^+^ central memory T-cell abundance in early adulthood among British-Pakistani participants, with differences from unaffected individuals diminishing by approximately age 50. No comparable pattern was observed among British-Bangladeshi participants (**Fig. 4d**). Similar variation by disease status, age and host background was observed across multiple immune populations (**Supplementary Fig. 23**), indicating that disease-associated immune differences vary across adulthood and demographic contexts.

Together, these analyses suggest that circulating immune composition reflects accumulated exposures and disease history through recurrent immune programmes shaped by age, sex and population context. The same diagnostic categories can therefore exhibit distinct immune profiles across cohorts with contrasting demographic and clinical characteristics, with implications for the generalisability of immune biomarkers and the design of immunomodulatory therapies.

### Cardiovascular therapies are associated with disease-related immune programmes

Pharmacological exposure may contribute to the disease-associated immune signatures described above. We therefore assessed whether these signatures were associated with commonly prescribed cardiovascular therapies and sought to identify treatment-associated changes in immune composition independent of underlying disease. We focused on statins and antihypertensive therapies, two widely used drug classes with established immunomodulatory properties. Analyses included 1,744 individuals without cancer or autoimmune disease and accounted for underlying disease, sex, age, BMI, smoking and relevant clinical covariates: total, LDL and HDL cholesterol for statins; systolic and diastolic blood pressure for antihypertensives. We first modelled associations between each drug exposure and scCBC abundance across the cohort, and then jointly modelled disease status, drug use and their interaction to distinguish disease- from treatment-associated effects and to test whether treatment modified the immune alterations associated with disease (**Methods**).

At conventional cell-type resolution (CT2), statin therapy showed limited association with immune-cell abundance. By contrast, cell-state resolved analyses (CT3) revealed reproducible associations within myeloid and T-cell compartments (**Fig. 5a, Supplementary Table 8**). Statin use was associated with reduced CD16^+^ monocytes together with lower number of differentiated CD8^+^ T cell and T follicular helper states, consistent with selective attenuation of inflammatory myeloid activation and adaptive immune differentiation, rather than broad changes in immune cell composition^53,54^. Antihypertensive therapy similarly yielded modest effects at the level of major immune lineages (e.g. classical monocyte and NK expansion with plasmablast depletion), but substantial heterogeneity across individual cell states (**Fig. 5a, Supplementary Tables 8**). Different antihypertensive classes were preferentially associated with distinct immune compartments, with significant associations for β- (ILC2 expansion) and α-blockers (naïve early B-cell expansion), and weaker associations for ACE (angiotensin-converting enzyme) inhibitors, ARBs (angiotensin II receptor blockers) and CCBs (calcium channel blockers; **Fig 5a**). These results indicate that pharmacological modulation is highly cell-state specific^55^ and qualitatively distinct from compositional signatures of disease reported earlier.

**Figure 5:**
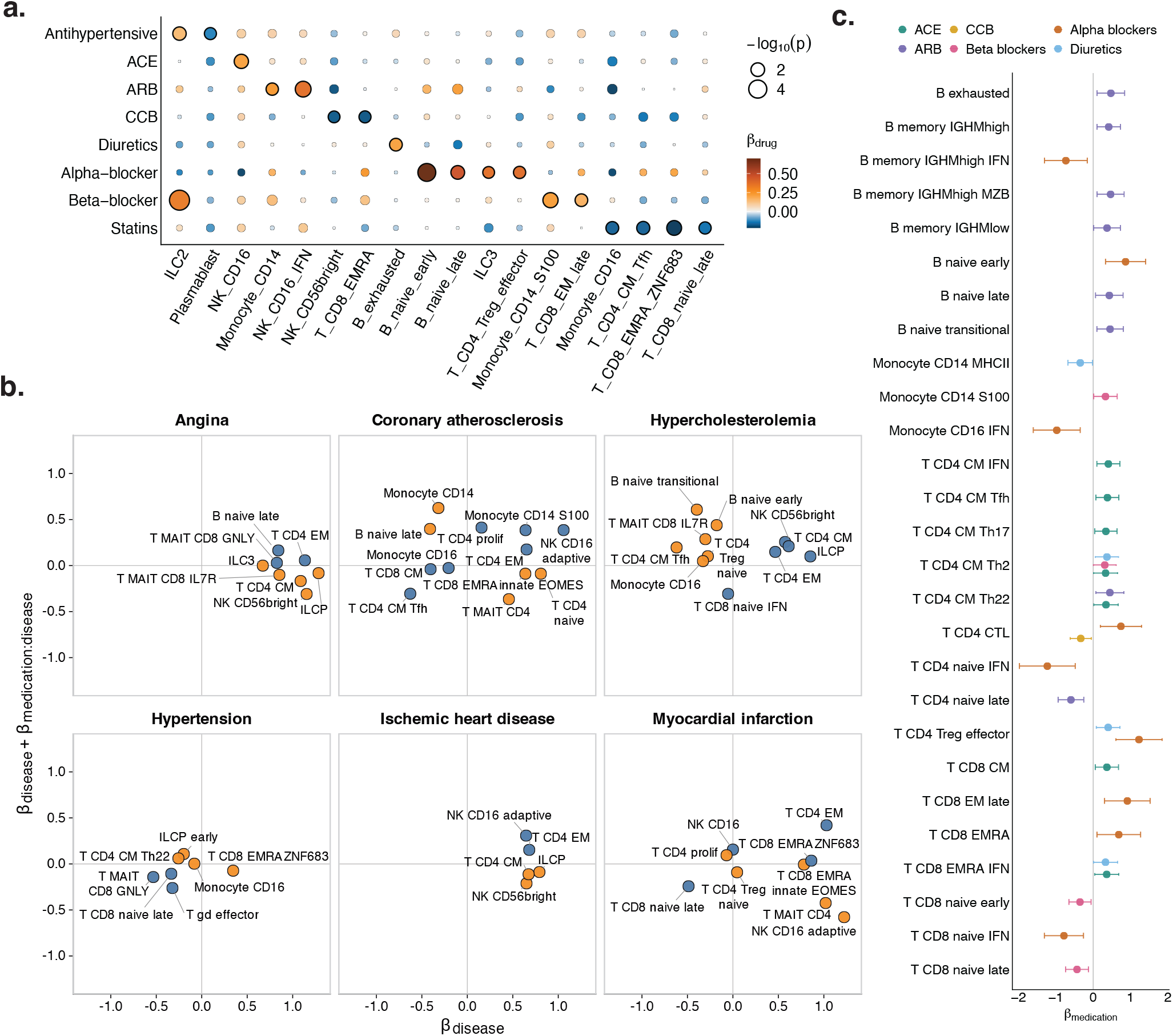
Associations between medication and scCBC traits. **a,** Drug–cell type dot plot showing associations between antihypertensive drugs, including subtypes, statins and scCBCs. Dot colour indicates the medication effect size (*β*) from generalised additive models, and dot size indicates significance (−*log*10(*p*)); black outlines denote associations with *p <* 0.05. Only cell types with at least one significant association are shown, ordered by the first significant drug. **b,** Statin–disease interaction scatterplot showing the associations between disease and cell-type abundance in untreated donors (*β_disease_*) and corresponding associations in statin-treated donors (*β_disease_* + *β_disease_*:*_medication_*). Points are coloured according to concordant or discordant direction of the disease effect, and labels indicate cell types. A shift toward zero on the y-axis relative to the x-axis indicates attenuation of the disease-associated effect in statin-treated donors, consistent with a shift toward control levels. **c,** Forest plot of antihypertensive effects on cell type abundance in hypertensive donors. Points and horizontal lines show effect sizes and 95% confidence intervals for significant associations. (ACE = angiotensin-converting enzyme inhibitors; ARB = angiotensin receptor blockers; CCB = calcium channel blockers)

To further test whether observational drug associations reflect the diseases for which treatments are prescribed rather than direct pharmacological effects, we next combined propensity-score matching (PSM) with generalised additive models (GAM) to distinguish treatment-associated immune remodeling from disease-associated immune signatures (**Methods; Supplementary Fig. 24**). Across cardiometabolic diseases, statins use was associated with attenuation of disease-related immune alterations (**Fig. 5b, Supplementary Tables 10**). Among all 33 disease–cell-type pairs showing nominal evidence of a disease association (p < 0.05) across 6 cardiometabolic conditions in UKB, the disease effect was smaller in absolute magnitude among statin-treated individuals, with 18 associations attenuated and 15 reversed effect while remaining closer to zero. Expansion of innate lymphoid progenitors observed in angina, ischaemic heart disease and hypercholesterolaemia was reduced among statin-treated individuals (disease × statin interaction β = −1.37, −0.88, and −0.75, respectively; all p < 0.01; **Fig. 5b**). Likewise, statin use was associated with reduced expansion of innate-like cell states in angina, including NK CD56^bright^ cells, memory CD4^+^ T cells and IL7R^+^ CD8^+^ MAIT cells (disease × statin interaction β = −1.46, −1.26, and −0.95, respectively; all p < 0.015). The largest effect was observed for NK CD16^+^ adaptive cells in myocardial infarction, where disease state was associated with a 1.22 s.d. higher counts in untreated donors, whereas the corresponding disease-associated effect among statin-treated donors was −0.58 s.d. (disease × statin interaction β = −1.80, p = 2.54 x 10^−4^). Other examples were CD4^+^ MAIT cells in myocardial infarction (disease × statin interaction β = −1.44, p = 5.37 x 10^−3^), NK CD56^bright^ and memory CD4^+^ T cells in ischaemic heart disease (disease × statin interaction β = −0.86 and −0.78, p = 8.41 x 10^−3^ and 0.019, respectively). Together, these findings indicate that statin use is associated with attenuation of cardiometabolic disease-associated immune alterations towards profiles observed in unaffected individuals, consistent with partial normalisation of immune composition rather than a uniform treatment-associated shift^43,54,56^.

Antihypertensive therapies likewise modified disease-associated immune states, although the affected populations differed across drug classes (**Fig. 5c**; **Supplementary Tables 9-12)**. ACE inhibitors, angiotensin receptor blockers, calcium-channel blockers, α-blockers, β-blockers and diuretics were each associated with distinct combinations of T-cell, B-cell, NK-cell and monocyte states within hypertensive donors^55,57^. Across all drug classes, 33 treatments–disease–cell-state associations met p < 0.05, involving 27 immune states. The strongest class-specific association was observed for α-blockers and CD4^+^ Treg effector cells (β = 1.23, p = 7.89 x 10^−4^), with others including naive late CD4^+^ T cells and ARBs (β = −0.59, p = 9.92 x 10^−4^), B naive early and α-blockers (β = 0.87, p = 4.65 x 10^−3^), naive late CD8^+^ T cells and β-blockers (β = −0.43, p = 6.28 x 10^−3^), and CD4^+^ central-memory IFN cells and ACE inhibitors (β = 0.41, p = 0.011). Overall, our results show that circulating immune composition reflects both disease status and treatment exposure, and that these signatures are distinct. These findings show that scCBCs can detect treatment-associated immune signatures in population cohorts and provide a basis for future longitudinal studies of pharmacological response.

### Single-cell immune composition improves discrimination of common diseases

Given that immune composition reflects the cumulative effects of ageing, environmental exposures and disease, we next assessed its capacity to distinguish individuals with and without common diseases and to identify the immune-cell populations contributing to this discrimination. We trained penalised logistic regression models to distinguish individuals with and without approximately 80 common diseases using CT2 scCBCs alone, estimating discriminative performance by repeated five-fold cross-validation (**Methods**). We then added demographic and lifestyle covariates (age, sex, ancestry, BMI and smoking status) to assess whether immune composition provided additional discriminatory information.

Immune composition alone provided moderate discrimination for multiple chronic diseases across both cohorts (mean AUCs ∼0.6–0.8), with the strongest performance observed for cardiometabolic diseases, demonstrating that disease-related information could be recovered from scCBCs without prior knowledge of demographic or lifestyle characteristics (**Fig. 6b, Supplementary Table 13**). Across all evaluated diseases, the mean AUC was 0.55 in UKB and 0.57 in G& H, and 13 of 82 and 12 of 40 conditions, respectively, achieved a mean AUC greater than 0.6. In UKB, coronary atherosclerosis, ischaemic heart disease, myocardial infarction, type 2 diabetes, obesity, hypercholesterolaemia and hypertension were consistently among the conditions with the highest discriminative performance. Conversely, acute infectious diseases showed little discriminatory signal, likely due to their transient nature (**Supplementary Table 13**). Similar patterns were observed in G& H, where type 2 diabetes, hypercholesterolaemia and hypertension achieved the highest discriminative performance, while musculoskeletal disorders, including chronic joint pain and Dupuytren disease, were also classified with AUCs above 0.6 (**Fig. 6c, Supplementary Table 13**). Of note, these signatures were not shared with obesity, potentially indicating that these are unlikely to capture obesity-induced joint pain or diabetes-associated Dupuytren disease.

**Figure 6:**
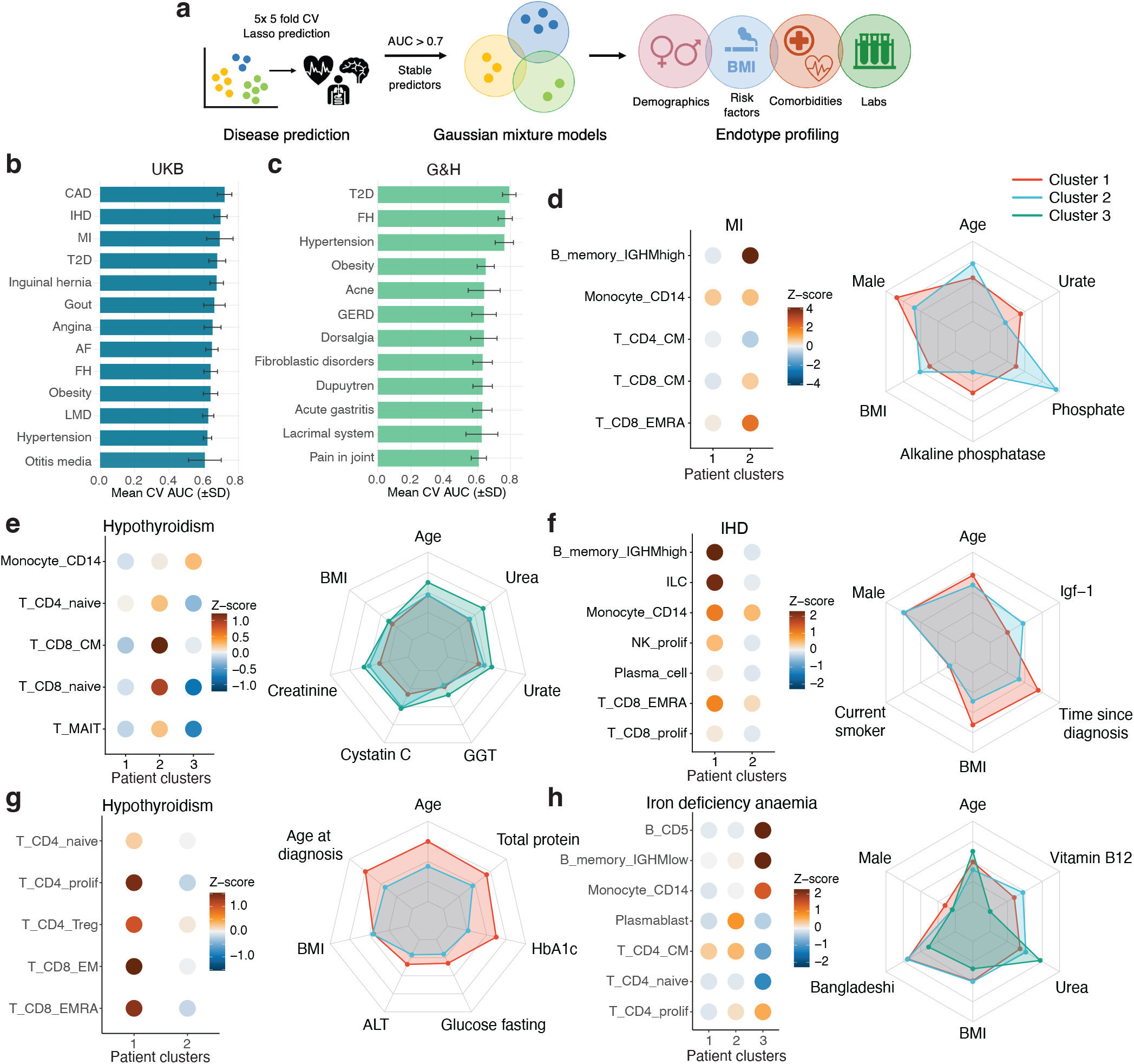
Disease discrimination and immune-defined patient subgroups. **a,** Schematic of the analysis pipeline for disease discrimination and identification of immune-defined patient clusters. **b-c,** Mean cross-validation AUC values, with standard deviation, for models using only scCBCs as input features, shown for diseases with mean AUC *>* 0.6 in UK Biobank (**b**) and Genes & Health (**c**). **d–h,** Dot plots of z-scores for scCBCs comparing cell types across immune-defined clusters, with corresponding radar plots showing differences in risk factors and comorbidities across clusters: myocardial infarction in UK Biobank (**d**), hypothyroidism in UK Biobank (**e**), ischaemic heart disease in UK Biobank (**f** ), hypothyroidism in Genes & Health (**g**) and iron deficiency anaemia in Genes & Health (**h**). Clusters are shown in red (1), blue (2) and green (3).

We next examined the immune features driving discrimination, rather than focusing solely on overall discriminative performance. The contributing cell populations broadly recapitulated the disease-associated programmes identified above. Across both cohorts, cardiometabolic diseases were characterised by expansion of inflammatory monocyte populations and depletion of naive T cells and enrichment of differentiated memory and effector lymphocyte compartments, whereas G& H additionally exhibited stronger CD4^+^ central memory signatures and reduced NK abundance (**Supplementary Fig. 25, Supplementary Table 13**). Together, these patterns suggest that disease discrimination was supported by coordinated changes across multiple immune populations rather than by any single cell type.

Finally, we assessed immune-derived discrimination alongside conventional demographic and lifestyle covariates. As expected, adding age, sex, ancestry, BMI and smoking status improved overall discriminative performance (**Supplementary Fig. 26, Supplementary Table 14**). Joint models achieved AUCs approaching or exceeding 0.8 for obesity, breast cancer and coronary atherosclerosis in UKB, and for obesity, dupuytren/fibroblastic disease, type 2 diabetes and hypercholesterolaemia in G& H. Across diseases, mean AUCs reached 0.62 in UKB and 0.65 in G& H, with gains exceeding 0.1 for 20 and 14 conditions, respectively. Inflammatory CD4^+^ monocytes in vascular disease and memory T-cell populations in metabolic disorders retained disease associations in these models, suggesting that immune-derived discrimination was not fully explained by demographic and lifestyle differences between cases and controls.

These analyses therefore show that high-resolution immune composition captures disease-discriminative information beyond conventional demographic and lifestyle characteristics, particularly for cardiometabolic and musculoskeletal conditions. scCBCs provide interpretable quantitative profiles reflecting the combined influences of ageing, environmental exposures and disease across distinct population contexts.

### Single-cell data immune composition resolves disease heterogeneity beyond clinical diagnosis

We next investigated whether high-resolution immune composition could resolve biologically distinct patient subgroups within diseases. For conditions with mean scCBC-based AUC > 0.7, we applied Gaussian mixture models to scCBCs to identify patient clusters, selecting the number of groups using the Bayesian information criterion. Clusters were subsequently characterised using donor variables (e.g. age, sex, ancestry, time to diagnosis), comorbidity profiles and quantitative disease biomarkers (**Methods**).

Across both cohorts, scCBC-based clustering identified clinically distinct subgroups within several common diseases (**Fig. 6d-f**). These clusters differed not only in immune-cell architecture but also in metabolic status, biochemical markers and comorbidity burden, indicating that immune composition captures clinically meaningful heterogeneity beyond disease diagnosis alone. Cardiovascular conditions in UKB shared recurrent inflammatory subgroups characterised by expansion of CD14^+^ monocytes and distinct lymphoid profiles (**Fig. 6d-f, Supplementary Fig. 27a**). Patients in these inflammatory clusters had greater adiposity, more pronounced dyslipidaemia and longer disease duration, with clinical biomarkers further delineating distinct metabolic and vascular profiles^58^. Across angina, ischaemic heart disease and myocardial infarction, increased CD14^+^ monocyte abundance was a shared disease feature, whereas distinct B- and T-cell profiles differentiated patient subgroups, consistent with immune diversification over the course of disease^59^. Hypothyroidism likewise comprised immune-defined subgroups with distinct clinical profiles (**Fig. 6e**). One subgroup showed increased inflammatory monocyte abundance and depletion of naïve CD8^+^ T and MAIT cells, accompanied by adverse renal, hepatic and metabolic biomarker profiles. A second subgroup had relatively preserved naïve and central memory T-cell populations and less pronounced biochemical abnormalities. These findings reveal substantial immune and metabolic heterogeneity among individuals sharing the same diagnosis.

Comparable heterogeneity emerged in G& H. Among patients with iron-deficiency anaemia, a small subgroup (8/153; 5.2%) showed increased CD5^+^ B-cell abundance -a population associated with potential pre-leukaemic profiles in our dataset (Giacopuzzi et al.)- alongside lower vitamin B12 (mean 296.9 versus 486.4 and 542.1 in the other clusters; FDR-adjusted p=0.031 and 0.019, respectively) and higher urea (5.32 versus 4.04 and 3.59; FDR-adjusted p=0.037 for both comparisons; **Fig. 6g, Supplementary Fig. 27b**). This subgroup links a distinct immune profile to concomitantly lower vitamin B12 levels, potentially reflecting combined effects of iron and vitamin B12 insufficiency on haematopoiesis^60^. In hypothyroidism, the smaller subgroup (17/75 patients; 22.7%) was older (49.5 versus 41.3 years; FDR-adjusted p = 0.020) and diagnosed later (43.9 versus 36.2 years; FDR-adjusted p = 0.024), with higher ALT (24.4 versus 21.3), fasting glucose (6.01 versus 5.32), HbA1c (48.3 versus 43.6) and total protein (75.6 versus 72.6; all FDR-adjusted p < 0.05; **Fig. 6h, Supplementary Fig. 27c**). This subgroup exhibited a less favourable metabolic and hepatic biomarker profile despite the absence of diagnosed diabetes^61^, suggesting that age and metabolic context contribute to immune heterogeneity in thyroid dysfunction, although subgroup profiles differed between cohorts. Overall, these analyses show that single-cell immune profiling reveals disease heterogeneity beyond conventional diagnostic categories. scCBCs delineate immune-defined subgroups with distinct metabolic and clinical profiles, supporting their potential for patient stratification and providing a basis for prospective evaluation of progression, recurrence and organ complications.

## Discussion

The immune system integrates genetic background with the cumulative effects of ageing, infection, metabolism and environmental exposures, yet these influences have traditionally been studied in isolation^4,32^. Here we combine population-scale single-cell transcriptomics with matched haematology measurements and electronic health records to quantify immune-cell composition across 5,079 individuals from two population cohorts spanning white European, British-Pakistani and British-Bangladeshi ancestry. Previous population studies established that human immune variation is strongly influenced by non-heritable factors and that age, shared environment and exposures such as smoking contribute substantially to variation in circulating immune-cell populations^26,32,62^. We extend this framework to single-cell resolution, showing how age, sex, lifestyle, disease and treatment jointly shape transcriptionally defined immune populations, and that immune composition carries informative signals for disease discrimination and for resolving immunologically and clinically distinct subgroups within the same diagnosis. By deriving quantitative single-cell blood counts, we generalise the clinical complete blood count to high-resolution immune states and establish immune composition as an integrative phenotype linking life-course exposures, population context and disease heterogeneity.

Large population single-cell studies such as OneK1K demonstrated the power of single-cell genomics for resolving inter-individual immune variation and its genetic determinants^15^. A central advance of our work is the development of quantitative single-cell immune phenotypes that overcome an important limitation of conventional single-cell compositional analyses^63^. Relative cell proportions are intrinsically interdependent, making it impossible to distinguish true biological expansion of individual populations from secondary changes driven by fluctuations elsewhere in the immune compartment. Anchoring single-cell proportions to matched clinical haematology measurements enables estimation of absolute immune-cell abundance while preserving the transcriptional resolution afforded by single-cell genomics. More broadly, this framework illustrates how established clinical phenotypes and high-dimensional molecular measurements can be integrated to generate quantitative traits suitable for population-scale analyses.

Our analyses reveal marked restructuring of immune composition across the adult life course. Consistent with previous studies^20–23^, ageing was characterised by contraction of naïve lymphocyte compartments together with expansion of differentiated cytotoxic and inflammatory myeloid populations, reflecting progressive immunosenescence. The broad age range of Genes & Health allowed us to resolve distinct, often non-linear, age-associated trajectories across immune populations that are difficult to capture in cohorts restricted to older adults. Sex effects were similarly dynamic, with several adaptive immune differences emerging only after midlife, indicating that immune sexual dimorphism develops progressively across adulthood^28^. Extending previous evidence that immune ageing can be non-linear^21,62^, our findings show that age-associated immune variation is neither uniform across cell types nor shared identically across populations, but is shaped by both sex and population context.

Among modifiable exposures, smoking and BMI had pronounced effects on immune composition. Importantly, both exposures shifted the immune system towards an older state beyond that expected from chronological age, consistent with accelerated immune ageing^7,34^. Smoking predominantly remodelled adaptive immune compartments, with effects that were largely reversible following smoking cessation, whereas adiposity produced broader alterations across innate and adaptive immunity that became increasingly pronounced with age. Notably, the attenuation of smoking-associated compositional changes after cessation contrasts with the persistent effects of prior smoking reported for adaptive immune responses and epigenetic state^7^, potentially suggesting a different rate of immune system recovery for cell composition compared to other functions of of the immune system, or other unmeasured effects not considered here. The data further enhances the fine-grained resolution of cellular compositional changes reported in previous studies. For instance, the observed expansion of early and late naïve B cells, CD4^+^ central memory subsets spanning Tfh, Th1, Th2, Th17 and Th22 lineages, and naïve Tregs, extends prior observations of obesity-linked broad B-cell and T-cell remodelling by resolving the specific cell states involved^34^. Similarly, UKB revealed BMI-associated decreases in NK CD16^+^ IFN^+^ and GNLY^+^/IL7R^+^ MAIT subsets, providing finer cell-state resolution relative to earlier reports of obesity-associated changes in NK and MAIT compartments^39^ and suggesting a more selective erosion of innate-like immune surveillance^40^.

A core finding of this work is that population context fundamentally shapes immune composition in health and disease. Even for shared diseases, immune programmes differed markedly between cohorts, with UK Biobank showing greater myeloid and cytotoxic remodelling and Genes & Health greater adaptive-memory, humoral and interferon responses. These differences likely reflect variation in age, multimorbidity, infectious exposure and disease prevalence, underscoring that immune signatures are not universal biomarkers but context-dependent products of pathology and population history^64^. At the same time, multiple clinically distinct diseases converged on a restricted set of conserved immune programmes. Rather than generating disease-specific immune signatures, cardiometabolic, inflammatory and infectious disorders repeatedly recruited shared modules involving inflammatory myeloid activation, adaptive-memory remodelling and type I interferon responses. This convergence suggests that diverse diseases exploit common components of immune organisation, with disease specificity arising primarily through differences in the relative contribution and timing of these programmes^65^. Other examples were the selective ILCP enrichment in digestive disorders (cholelithiasis, gastro-oesophageal reflux disease, other gastric–duodenal conditions) suggesting a shared precursor-like innate lymphoid programme across the upper gastrointestinal tract, potentially reflecting activation at mucosal barriers preceding differentiation into effector subsets for tissue repair and immune regulation^66^. Similarly, humoral remodelling was prominent in skin cancers and dermatitis (infective and seborrhoeic), with CD5^+^ B-cell enrichment. Overall, our model provides a framework for understanding the substantial overlap between genetic susceptibility, environmental exposures and chronic inflammatory diseases, and supports the concept that immune composition represents an intermediate phenotype through which multiple pathogenic processes are integrated^1–3,10,11^.

The availability of electronic health records further enabled immune composition to be studied within its broader clinical context. Medication analyses demonstrated that cardiovascular therapies partially attenuated disease-associated immune states, highlighting the importance of accounting for treatment exposure when interpreting immune variation in observational studies. While our study size allowed explorations limited to the most-commonly used drugs, our data suggests treatment effects to be distinct from disease signatures. More resolution will come with larger, controlled experiments. For instance, the association of CD14^+^ monocytes and coronary atherosclerosis seems to increase in statin-treated patients, but only the disease-drug effect was significant, while the disease term was not. This may reflect limited power to detect the disease association among untreated individuals, or treatment status may capture differences in disease severity or timing, for example if individuals with more severe coronary atherosclerosis were more likely to receive treatment or initiated treatment after disease onset.

Beyond association mapping, high-resolution immune composition contributed information relevant to the discrimination of several common chronic diseases and identified immune-defined subgroups of patients sharing the same diagnosis but differing in metabolic characteristics and comorbidity burden. That this degree of biological and clinical structure can be recovered from a single cross-sectional measurement highlights the potential of prospective longitudinal immune profiling in population cohorts. Repeated sampling could move beyond between-individual associations to capture within-individual immune dynamics, establish how immune states evolve before and after disease onset or treatment, and identify dynamic biomarkers of progression and therapeutic response. Although the unprecedented resolution achieved in this work is beyond what can be implemented in the clinical setting, these results demonstrate that transcriptionally resolved immune composition captures clinically relevant biological information beyond conventional demographic and lifestyle variables. As population-scale single-cell datasets continue to expand and longitudinal sampling becomes increasingly feasible^67^, immune composition may become a useful quantitative biomarker for patient stratification, disease monitoring and therapeutic response.

The inherent limiting features of this study point to opportunities for future work. While scCBCs substantially improve the interpretability of single-cell compositional data by providing estimates of absolute immune-cell abundance, these remain inferred from matched haematological measurements rather than directly measured for each transcriptionally defined population. Furthermore, our analyses were restricted to peripheral blood mononuclear cells and therefore do not capture granulocyte populations or tissue-resident immune compartments that contribute to many diseases. The integration of electronic health records enabled immune composition to be studied across a broad range of diseases, exposures and treatments, although the observational design does not by itself establish causality and residual confounding from factors such as disease severity, treatment adherence or unmeasured exposures may remain. Finally, while the inclusion of two ancestrally and demographically distinct cohorts represents a major strength of this study, extending similar analyses to additional global populations will be essential for capturing the full spectrum of human immune diversity and local exposures.

In summary, quantitative single-cell blood counts reveal a structured landscape of human immune variation shaped by ageing, sex, modifiable exposures, disease and treatment. Common diseases repeatedly perturb a restricted set of shared immune programmes, but the expression of these programmes varies substantially across population contexts and captures clinically meaningful heterogeneity within individual diagnoses. As population-scale molecular cohorts become increasingly linked to longitudinal health records, this framework provides a foundation for dissecting how these influences combine to shape human immune variation and disease.

## Materials and methods

### Ethics

Recruitment descriptions: UK Biobank obtained approval from the North West Multi-centre Research Ethics Committee (MREC) UK NHS REC approval reference number: 21/NW/0157. Genes & Health obtained ethical approval from London South East NRES Committee of the Health Research Authority approval reference number: 14/LO/1240.

### The CARDINAL dataset

Generation of the CARDINAL scRNA-seq dataset, including sample collection, data preprocessing and cell-type annotation, is described in the accompanying manuscript by Giacopuzzi et al. Complete blood counts were measured from matched blood samples using a Sysmex XN-1000 haematology analyser, as detailed in the same manuscript.

### Harmonising Sysmex and 10x measurements to define high-confidence samples

Data points identified as outliers based on discrepancies between lymphocyte-to-monocyte ratios from Sysmex and 10x measurements were excluded, ensuring consistency between technologies and reducing potential measurement error.

The quality-controlled dataset used for downstream analyses included only fresh samples, as 10x is known to be less reliable for monocyte quantification in frozen material, and their inclusion could introduce bias.

Outliers were defined using an interquartile range (IQR)-based criterion: a sample was removed if

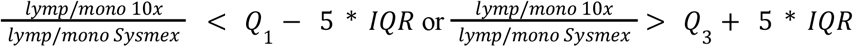

where Q_1_, Q_3_ and IQR denote the first quartile, third quartile and interquartile range of the ratio distribution, respectively. We chose a threshold of 5 IQR to avoid removing an excessive number of data points.

After filtering, the correlation between Sysmex and 10x lymphocyte-to-monocyte ratios was 0.68 for UKB and 0.54 for G& H. When fitting a regression model without an intercept, the correlation coefficients were R = 0.86 for UKB and R = 0.84 for G& H (**Supplementary Fig. 2**). The final cohorts used for the analyses are reported in **Supplementary Table 1**.

### Definition of single-cell complete blood counts (scCBCs)

After outlier removal, single-cell complete blood counts (scCBCs) were derived by integrating data from the Sysmex haematological analyser with 10x single-cell profiles. Total lymphocyte and monocyte counts were first extracted from Sysmex complete blood counts, and the proportion of each cell type within the lymphocyte and monocyte compartments was then computed from the 10x data.

For each cell type, scCBCs were obtained as:

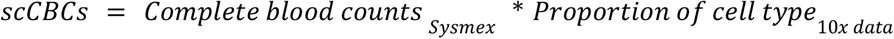

with this procedure applied separately for lymphocytes and monocytes. The same procedure has been used to compute scCBCs for CT2 and CT3. The resulting distributions of scCBCs across cell types are shown in **Supplementary Fig. 1**.

This integration provides estimates of absolute immune-cell abundance rather than proportions alone, enabling quantitative comparisons across individuals and cohorts. Although the 10x proportions used to derive scCBCs remain relative within the lymphocyte or monocyte compartment, scaling them by each donor’s measured compartment count means that the resulting scCBCs are not constrained to the same total across individuals. Thus, cell subsets within each compartment remain partly related through their shared compartment count, but the compositional constraints and associated correlation structure observed when analysing proportions alone are substantially reduced.

### Associations with biological covariates and risk factors

We considered age, sex, ancestry, smoking status, body mass index (BMI), and the index of multiple deprivation (IMD) as potential covariates. Age was computed at the time of PBMC collection, while smoking status and BMI were assigned based on the latest available measurement. The index of multiple deprivation incorporates several socioeconomic factors, including access to services, community safety, crime, education, employment, health, housing, income, living environment, and physical environment scores. Since the indices are calculated in different ways between England, Scotland and Wales, and are provided separately by UKB, we first standardized them within each country to ensure comparability before merging them into a single variable. In G& H it is available for North East London patients only (n = 772). Ancestry was determined using the Principal component Ancestry proportions using non-negative least squares Estimation (PANE) method^68^. Due to data availability, in G& H smoking status and BMI were available for only a subset of individuals (n = 773 and 770, respectively).

For each cell type, we performed a linear model to assess associations between biological factors and cell-type abundances. The models were specified as follows:

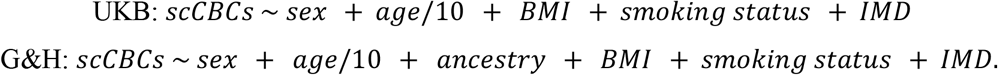

To ensure comparability across cell types, scCBCs were variance-scaled, while preserving their original mean to retain biologically meaningful differences in absolute abundance. Continuous covariates (BMI and index of multiple deprivation) were standardised, whereas categorical variables (sex, ancestry, and smoking status) were included either as binary or multi-level factors. Age was not standardised to maintain interpretability in years.

Accordingly, beta coefficients represent the expected change in standardised cell abundance associated with a one-unit increase in a continuous covariate or with membership of a given category relative to the reference level. Positive and negative coefficients therefore indicate higher and lower abundance, respectively. We used linear models to prioritise directionality and consistency of association across cell types rather than exact effect size estimation. Multiple testing was controlled using the Benjamini–Hochberg procedure. Since many cell types exhibited non-normal distributions, we explored various transformations. However, log and gamma transformations were unsuitable due to the high prevalence of zeros, and alternative transformations did not meaningfully improve normality or model estimation. To account for multiple testing, we applied the Benjamini-Hochberg correction to the p-values.

### Age-associated variation in immune-cell composition

To characterise age-associated variation in immune-cell composition, scCBC abundances were modelled as smooth functions of age using functional data analysis (FDA). Before smoothing, scCBCs were adjusted by linear regression for the covariates included in the corresponding association models, excluding age and, for stratified analyses, the covariate defining the groups under comparison. Smoothing was applied to the resulting residuals using five B-spline basis functions and penalised least squares with a second-derivative roughness penalty (λ = 1 × 10⁻⁴). For visualisation, fitted curves were evaluated across ages 53–85 years in UKB and 16–89 years in G& H. Pointwise 95% confidence intervals were calculated as: *f(x) ± 1.96 × SE(x)*, where *f(x)* denotes the fitted value at age *x*. All FDA analyses were implemented using the *fda* package in R^69^.

For compact visualisation of between-group differences, age-specific contrasts were calculated as:

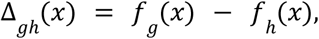

where *f_g_(x)* and *f_h_(x)* are the fitted curves for the comparison and reference groups, respectively. Female donors were contrasted with male donors, and donors with overweight or obesity with those of healthy weight. Contrast profiles were standardised within each cell type across age, and cell types were ordered by average-linkage hierarchical clustering using *1 − r*, where *r* is the Pearson correlation between standardised profiles.

In G& H, we additionally tested whether age-associated patterns deviated from linearity. scCBCs were scaled within each cell type by their standard deviation and BMI was standardised before covariate adjustment. For each cell type, abundances were adjusted for sex, ancestry, BMI and smoking status and the resulting residuals were fitted using both a linear age model and the FDA model. Evidence for non-linearity was quantified as:

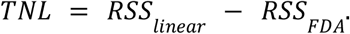

Significance was assessed using 5,000 wild-bootstrap replicates generated under a linear null model containing age and all adjustment covariates, with residuals independently multiplied by −1 or 1. Empirical p-values were adjusted across cell types using the Benjamini–Hochberg procedure. Cell types with fewer than 30 complete observations or fewer than seven unique ages were excluded. Fitted curves and their first and second derivatives were evaluated over 200 equally spaced age points.

We next tested whether age-associated patterns differed by sex, ancestry, BMI or smoking status in G& H. BMI was classified as healthy weight (18.5–<25 kg/m²), overweight (25–<30 kg/m²) or obesity (≥30 kg/m²); underweight donors were excluded. Smoking status was classified as never, previous or current. Analyses were restricted to complete observations and to comparisons in which each subgroup contained at least 20 donors. Subgroup-specific FDA curves were fitted after adjustment for the remaining covariates, and pairwise comparisons were restricted to the overlap between the 2.5th and 97.5th percentiles of the subgroup age distributions. For groups *g* and *h*, curve differences were quantified by the mean integrated squared difference:

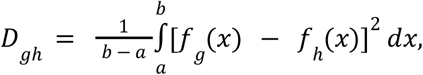

where *[a, b]* denotes their shared age support. For modifiers with more than two levels, the overall statistic was the sum of *D_gh_*across pairwise comparisons.

Overall and pairwise curve-equality p-values were estimated from 5,000 wild-bootstrap replicates generated under the null hypothesis of a common age-associated curve across subgroups. Overall p-values were adjusted using the Benjamini–Hochberg procedure across cell types within each modifier and across all modifier–cell-type tests, whereas pairwise p-values were adjusted using the Holm procedure. To localise differences along the age range, 95% simultaneous confidence bands were constructed from a separate set of 5,000 wild-bootstrap replicates. Bands were defined as:

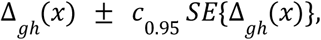

where *c_0.95_* was the 95th percentile of the maximum absolute standardised bootstrap deviation across ages and contrasts. Contiguous age points for which the simultaneous band excluded zero were combined into supported age intervals. Because these intervals were defined on a finite 200-point age grid, their boundaries were interpreted as approximate.

### Partial least squares regression to assess determinants of cellular ageing

Partial least squares (PLS) regression was performed in R (*pls* package^70^) to derive a latent cellular ageing axis from scCBCs measured at CT2 level. Cell type abundances were assembled into a predictor matrix, mean-centred and scaled to unit variance before model fitting. PLS identifies latent components as linear combinations of the predictors that maximise covariance with the response variable; here, chronological age was used as the response. The model was fitted with up to 10 components and evaluated by cross-validation, and PLS1 was retained as the primary ageing axis because it captured the strongest age-aligned signal. Loadings were examined to identify the cell types contributing most strongly to this axis. To quantify how risk factors modified the ageing score, PLS1 was modelled as a function of age, sex, ancestry, BMI and smoking status:

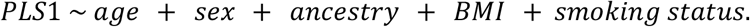

For visualisation, age acceleration was defined as the residual from regressing PLS1 on chronological age, with positive values indicating accelerated ageing and negative values indicating decelerated ageing. Regression estimates are reported with 95% confidence intervals.

### Stability of non-genetic covariates over time

Smoking behaviour showed limited variability over follow-up. Between baseline assessment and PBMC collection, 245 individuals reported quitting smoking, whereas 39 reported initiating smoking.

BMI showed greater temporal variability, with 1,043 participants changing BMI category during follow-up. We therefore assessed the robustness of BMI-related associations using four alternative measures: BMI at baseline, BMI at recall, the polygenic risk score (PRS) for BMI, and clinical diagnoses of obesity. BMI at baseline refers to measurements collected between 2006 and 2010, while BMI at recall corresponds to the most recent measurement available from the imaging visits conducted from 2014 onwards. The PRS for BMI was computed by UK Biobank (Field 26216). Obesity diagnoses were extracted from electronic health records as described in methods; it is important to note that only the first recorded diagnosis was available, and thus we could not ascertain whether individuals subsequently reversed their obesity status.

To evaluate whether these covariates had consistent effects on immune cell-type abundance, we performed four separate linear models, one for each covariate:

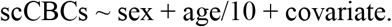

scCBCs were variance-scaled to ensure comparability across cell types, while preserving their original mean to retain biologically meaningful differences in absolute abundance. Continuous covariates were standardised, whereas categorical variables were included as binary factors. Age was not standardised to maintain interpretability in years.

### Disease definition

We derived 330 disease definitions from the MVP–Finngen–UKB meta-analysis phecode set (https://mvp.finngen.fi/), manually supplementing with 20 additional autoimmune and mental health disorders to enhance coverage. Three were subsequently excluded due to duplication (essential hypertension) or challenges in extracting reliable diagnoses (glomerular disorders classified elsewhere and kidney transplant status).

Disease definitions corresponded to phecodes, which aggregate individual ICD codes into clinically meaningful categories, thereby reducing sparsity and enhancing statistical power in association analyses. Phecodes were mapped to ICD-10, Read2, and CTV3 codes using the UK Biobank Read-to-Phecode repository^71^. In UK Biobank, diagnoses were extracted from Hospital Episode Statistics (HES), cause-of-death records, cancer registries, and general practitioner (GP) clinical events using the *ukbrapR* package^72^. For Genes & Health, three- and four-digit ICD-10 codes were available, already curated by the cohort consortium, and also included SNOMED from primary care and other sources, where a SNOMED code could be mapped to a single ICD10. Diagnoses were extracted from their first recorded occurrence. Implausible dates (e.g. diagnosis pre-birth) were removed.

Further details, including the full list of diseases and case counts, are provided in **Supplementary Table 2**.

### Associations between immune cell types and diseases

Associations between single-cell complete blood counts (scCBCs) and disease status were assessed using generalised additive models (GAMs), which allow non-linear associations between covariates and immune cell abundances. Analyses were limited to diseases represented by at least 35 cases. Separate models were fitted for each cell type, as specified below:

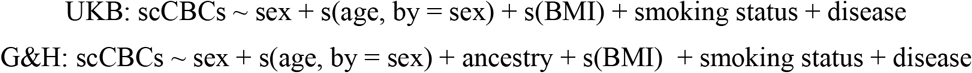

where *s*(⋅) denotes smooth functions estimated using B-spline basis expansions. Covariates were processed in accordance with prior analyses: age and BMI were standardised to z-scores, and sex, smoking, ancestry, and disease status were encoded as categorical variables. To ensure comparability across models, scCBCs were variance-scaled within cell types while preserving mean biological differences. Accordingly, beta coefficients for disease status represent the expected difference in standardised cell abundance between cases and controls, after adjustment for covariates, with positive values indicating higher abundance in cases and negative values indicating lower abundance. In G& H, smoking data and BMI information were available for *n*=773 and 770 participants respectively. Control cohorts were stringently defined as individuals without diagnoses belonging to the same disease category (e.g., controls for type 2 diabetes excluded all metabolic disorders), mitigating potential misclassification and class imbalance. Participants with malignant neoplasms were excluded from both case and control groups to isolate disease-specific immune variation from oncological effects. P-values for disease effects were adjusted for multiple testing using the Benjamini–Hochberg false discovery rate procedure. All models were fitted in *R* using the *mgcv* package^73^, with smoothness parameters estimated via restricted maximum likelihood (REML).

### Disease network

A disease–cell type signature network was constructed from UKB and G& H cohort-specific disease–cell type (CT3) association results. For each disease and cohort, we defined a signed cell-type signature by retaining cell types with nominal association at p<0.05 and encoding the direction of the disease effect (β≥0 as up, β<0 as down). Disease–cohort nodes were retained only if they contained at least two such signed cell types. Nodes represent disease–cohort pairs; their size reflects the number of nominal hits. Edges were drawn between two nodes when they shared at least two signed cell types and their Jaccard similarity (intersection over union) was at least 0.2. Isolates were removed for display.

The network was visualised using a stress layout. Node colour indicated disease category and node shape indicated cohort (circle for UKB, triangle for G& H). Labels were shown only for the most connected or largest nodes to maintain visual clarity. All steps were implemented in R using *igraph*, *tidygraph* and *ggraph*, with random seed 123 for reproducibility.

### Primary care prescription data pre-processing

Primary care prescription data were obtained from UK Biobank general practitioner (GP) records. Medications were identified using multiple coding systems, including Read v2, British National Formulary (BNF), and Dictionary of Medicines and Devices (dm+d) codes. The dataset also contained drug names, prescribed quantities, and issue dates. Records were restricted to participants with available linkage in the CARDINAL dataset. To improve the completeness of drug names, missing entries were imputed from Read v2 code descriptions using the lookup table provided by UK Biobank (Resource 592; https://biobank.ndph.ox.ac.uk/showcase/refer.cgi?id=592), which maps Read codes to canonical medication terms (67,614 code–term pairs). Drug names were harmonised by merging the dataset on Read v2 codes. Drug names were subsequently normalised to represent compound names only. All terms were converted to lowercase, and parenthetical expressions, dosage indicators, formulation descriptors (e.g., “tablets”, “capsules”, “cream”, “patch”), and numeric values were removed. Cleaned names were then converted to title case and whitespace-normalised to produce concise drug identifiers. To consolidate synonymous entries, a collapsing rule was applied to replace longer or derivative names with their shortest unique prefix, yielding a single canonical representation for each distinct medication. The final dataset comprised 283,053 prescriptions from 2,260 CARDINAL participants, corresponding to 2,265 unique cleaned drug names.

### Associations between immune cell types and drug prescriptions

We assessed the effect of commonly prescribed drugs, considered as comorbid exposures, on cell-type abundance, independently of the diseases for which they are typically prescribed. Specifically, we considered statins and antihypertensives, where statin exposure was defined as any prescription of atorvastatin, pravastatin, rosuvastatin, or simvastatin, while antihypertensive exposures comprised five classes: diuretics, calcium channel blockers (CCBs), ACE inhibitors, angiotensin receptor blockers (ARBs), β-blockers, and α-blockers. Medication prescription data were available for 1,840 donors. After excluding donors with cancer or autoimmune disease diagnoses, analyses were performed on 1,744 individuals. Cell-type abundance (scCBC) was modelled for each immune cell type and each drug using generalised additive models (GAMs) of the form:

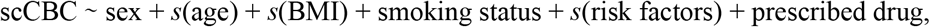

where smooth terms s(⋅) captured nonlinear effects of age, BMI and risk factors, and prescribed drug was a binary indicator for exposure to each specific medication (tested individually). Risk factors were selected individually for each drug to control for potential confounding due to differences in baseline levels of drug targets across donors. Specifically, HDL, LDL, and total cholesterol were included for statins, while diastolic and systolic blood pressure were included for antihypertensives. scCBCs were variance scaled, while all the numerical covariates were standardised.

### Disentangling disease status and drug prescription effects on cell type abundance

To disentangle disease-associated immune alterations from medication-related effects, we extended our previous statistical models by jointly modelling disease status, prescribed drug use, and their interaction. Medication prescription data were available for 1,840 donors. After excluding donors with cancer or autoimmune disease diagnoses, analyses were performed on 1,744 donors. Controls were further filtered to exclude individuals with related conditions (e.g., for myocardial infarction analyses, controls without myocardial infarction but with ischaemia were removed).

We implemented a two-stage pipeline combining propensity score matching (PSM) with generalised additive models (GAMs) to estimate associations between cardiovascular drug prescriptions and standardised immune cell-type abundances (scCBC), stratified by disease status. Statin exposure was defined as any prescription of atorvastatin, pravastatin, rosuvastatin, or simvastatin. Antihypertensive exposures comprised six classes: diuretics, calcium channel blockers, ACE inhibitors, angiotensin receptor blockers, β-blockers, and α-blockers. Disease indicators included hypercholesterolaemia (E4_HYPERCHOL), myocardial infarction (I9_MI_STRICT), ischaemic heart disease (I9_IHD), hypertension (I9_HYPTENS), coronary artery disease (I9_CORATHER), and angina pectoris (I9_ANGINA).

For drug–disease pairs permitting matched cohorts (statins across diseases; antihypertensives across all classes), PSM was conducted separately for each disease using 1:1 nearest-neighbour matching without replacement (*MatchIt package*). Propensity scores were estimated via logistic regression on age, sex, BMI, smoking status, disease indicator, and risk factors (for statins: LDL, HDL, total cholesterol; for antihypertensives: diastolic and systolic blood pressure). When healthy (disease-free) drug-exposed donors were unavailable, matching was restricted to diseased individuals, conditioning on age, sex, BMI, smoking, and risk factors. Covariate balance was evaluated using Love plots of absolute standardised mean differences (threshold 0.1; cobalt).

Matched samples underwent cell-type-specific standardisation (scCBC scaled by cell-type standard deviation). Associations were then modelled using GAMs (*mgcv package*):

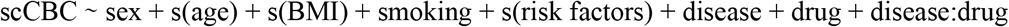

or, within diseased strata,

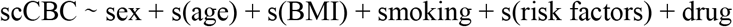

(cubic regression splines, bs=‘cr’). Parametric coefficients and smooth-term p-values were extracted for drug main effects and disease–drug interactions.

### Disease discrimination

Disease risk was modelled using penalised logistic regression with repeated cross–validation, followed by an unpenalised inference model. For each disease endpoint, we constructed a binary outcome (case vs control), and tested only with diseases at least 50 cases. Controls were stringently defined as individuals lacking any diagnosis within the same disease category (e.g., for type 2 diabetes, controls excluded all metabolic disorders), minimising misclassification and class imbalance. Participants with malignant cancers were excluded from both cases and controls to isolate disease-specific immune signatures from oncological confounds.

We fitted two complementary models for each endpoint: (i) a scCBC–only model, in which disease status was predicted using CT2 immune–cell abundance alone; and (ii) a combined model, in which the same scCBC features were augmented with established risk factors. For the combined model, continuous predictors were z–scaled. Only scCBCs at the CT2 level were used, to preserve biologically interpretable immune subsets while limiting overfitting.

To obtain robust disease discriminations, we performed stratified 5-fold cross-validation of an L1-penalised logistic regression (lasso) model. Within each training fold, we fitted a binomial lasso model using *glmnet*, optimising the penalty parameter via internal cross-validation with deviance as the loss function. To account for class imbalance, we incorporated inverse-frequency class weights to downweight the majority class. Out-of-fold predicted probabilities were generated for all individuals, and discrimination was quantified using the area under the receiver operating characteristic curve (AUC). To improve robustness and assess selection stability, we repeated the full 5-fold cross-validation procedure 5 times using independent random fold assignments. Predictors selected in at least 50% of runs were retained for downstream inference. For disease discrimination including risk factors, age, sex, smoking status and BMI were forced into the final model irrespective of penalised selection to ensure epidemiological interpretability.

For interpretability, we refitted an unpenalised multivariable logistic regression model on the full dataset, including only the stable predictors plus forced covariates, and obtained regression coefficients, standard errors, Wald z-statistics, p-values and odds ratios. Overall model performance was summarised by the distribution of AUCs across all folds and repetitions (n = 25).

### Gaussian mixture model endotyping

Disease endotypes were identified for diseases previously predicted with mean AUC > 0.7 (covariates + scCBC predictors), resulting in 18 diseases in UKB and 16 in Genes and Health. Only cases were considered; for each disease, cell-type predictors were pre-selected via lasso regression from prediction models, excluding intercept, age, sex, ancestry, smoking status, and BMI. Numeric cell-type abundance was z-score scaled.

Gaussian mixture models (GMMs; *mclust* package) with 1–4 components were fitted to the scaled cell-type matrix using ellipsoidal covariance structures (EEE: equal volume/shape/orientation; VVV: variable volume/shape/orientation), with optimal model selection by Bayesian Information Criterion. Cases received hard classifications (1 to *G*) via maximum *a posteriori* probability. Endotype assignments were linked to donor metadata (age, sex, ancestry, smoking status, BMI, cell profiles) for downstream analyses.

### Cluster profiling and statistical comparisons

Endotype clusters were characterised using a comprehensive non-parametric framework. Donor data (age, sex, ancestry, smoking status, BMI) were augmented with time since diagnosis, age at diagnosis, blood-based risk factors (n=32 in UKB: alanine aminotransferase, albumin, alkaline phosphatase, apolipoprotein a/b, aspartate aminotransferase, c reactive protein, calcium, hdl/ldl/total cholesterol, creatinine, cystatin c, direct/total bilirubin, gamma glutamyltransferase, glucose, glycated haemoglobin hba1c, igf-1, lipoprotein a, oestradiol, phosphate, rheumatoid factor, shbg, testosterone, total protein, triglycerides, urate, urea, Vitamin D, diastolic/systolic blood pressure; n=47 in G& H: Albumin, ALP, ALT, AST, Bilirubin, Blood ketones, C3, C4, Calcium, CK, creatinine, CRP, diastolic/systolic/mean arterial blood pressure, Ferritin, Folate, Fructosamine, GGT, fasting/non-fasting glucose, Haematocrit, Hb, glycated haemoglobin hba1c, hdl/ldl/total/non-hdl cholesterol, Heart_Rate, Insulin IgG, Iron saturation, Mg, MPO antibody, oestradiol, OGTT 120mins, Phosphate, PTH, Rheumatoid factor, T4 free, testosterone, Total protein, Triglycerides, TSH, Urate, Urea, Vitamin B12, Vitamin D total), medications (statins, metformin, antihypertensives; UKB only) and comorbidities (hypertension, T2D). Singleton clusters (n<2 donors) were excluded from hypothesis testing but aggregated (“SmallN”) for summary statistics. Inter-cluster differences were assessed via Kruskal–Wallis with Dunn post-hoc tests (Benjamini-Hochberg adjusted) for continuous covariates, and χ²/Fisher exact tests (simulated p-values, B=10⁴ if expected frequencies <5, cells <6, or total n<50) for categorical variables and comorbidities.

## Data Availability

All individual-level data used in this study are available through approved applications to UK Biobank and Genes & Health; summary results are provided in the manuscript and supplementary materials.

## Acknowledgements

This research was funded in part by the Wellcome Trust [grant number 220540/Z/20/A], the Chan Zuckerberg Foundation [grant number 2020-224316] and by the British Heart Foundation (BHF) Cambridge Centre for Research Excellence RE/18/1/34212. Fondazione Human Technopole is supported by public funds awarded by the Italian Ministry of Economy and Finance, the Ministry of University and Research, and the Ministry of Health.

This work is supported by the project IMPACT (G.A.N.) 101201772 Funded by the European Union. Views and opinions expressed are however those of the author(s) only and do not necessarily reflect those of the European Union or grant authority (ERCEA). Neither the European Union nor the granting authority can be held responsible for them.

This project has received funding from the Innovative Medicines Initiative 2 Joint. Undertaking (JU) under grant agreement No 101007873 (EU-IMI2 101007873).

L.A. was supported by the Helmholtz Association, as part of the joint research school Munich School for Data Science (MUDS) and the Studienstiftung des deutschen Volkes (German Academic Scholarship Foundation).

This work was supported by the Deutsche Forschungsgemeinschaft (DFG) - Projektnummer 452881907 (grant CRC TRR 338).

E.R and F.J.T are funded by the European Union ERC, DeepCell - 101054957.

This research has been conducted using the UK Biobank Resource under application number [69938].

We thank Wellcome Sanger Institute scientific operations team members; Stefanie Lensing, Scott Thurston, Lesley Shirley, Sameena Iqbal, Katy Taylor, Cordelia Langford and Ian Johnston for automation, sample management and sequencing operations support. Wellcome Sanger Institute Human Genetics Department team members; Sally Bygraves, Nicola Corton, Kalia Dede, Guillaume Noel and Hannes Postingl.

Genes & Health is/has recently been core-funded by Wellcome (WT102627, WT210561), the Medical Research Council (UK) (M009017, MR/X009777/1, MR/X009920/1), Higher Education Funding Council for England Catalyst, Barts Charity (845/1796), Health Data Research UK (for London substantive site), and research delivery support from the NHS National Institute for Health Research Clinical Research Network (North Thames). We acknowledge the support of the National Institute for Health and Care Research Barts Biomedical Research Centre (NIHR203330); a delivery partnership of Barts Health NHS Trust, Queen Mary University of London, St George’s University Hospitals NHS Foundation Trust and St George’s University of London. Genes & Health is/has recently been funded by Alnylam Pharmaceuticals, Genomics PLC; and a Life Sciences Industry Consortium of AstraZeneca PLC, Bristol-Myers Squibb Company, GlaxoSmithKline Research and Development Limited, Maze Therapeutics Inc, Merck Sharp & Dohme LLC, Novo Nordisk A/S, Pfizer Inc, Takeda Development Centre Americas Inc. We thank Social Action for Health, Centre of The Cell, members of our Community Advisory Group, and staff who have recruited and collected data from volunteers.

We thank: Barts Health NHS Trust, NHS Clinical Commissioning Groups (City and Hackney, Waltham Forest, Tower Hamlets, Newham, Redbridge, Havering, Barking and Dagenham), East London NHS Foundation Trust, Bradford Teaching Hospitals NHS Foundation Trust, Public Health England, Discovery Data Service/Endeavour Health Charitable Trust (especially David Stables), Voror Health Technologies Ltd, NHS England - for GDPR-compliant data sharing backed by individual written informed consent.

Most of all we thank all of the volunteers participating in UK Biobank and Genes & Health.

## Author contributions

G.C. contributed to project discussion and result interpretation. S.W. generated and helped interpret the Sysmex haematology data. O.D. performed cell-type annotation and contributed to its interpretation.

G.I., D.v.S., E.L.C., F. Torabi., M.H., N. Subasingha., L.D. and S.W. generated the data.

M.O., A.R., D.B., E.G., R.E., and V.I. performed data processing.

E.G., S.W., M.O., A.C., and A.R. carried out data quality control and benchmarking.

J.P.E., S.L., A.R., O.D. and F.C. conducted data integration, cell annotation and refinement.

A.R. performed ancestry definition analysis.

A.L., S.S., A.S., B.A., E.H., M.J.B. L.A., E.R., B.C., M.T., P.B., D.V., C.A.G.,C.C., M.I.C., I.G.,

C.D.C., M.C., F.S., A.G., V.S., J.G. assisted with interpretation of results.

S.W., N.Staudt., V.I., S.A.T., M.J.B., O.S., D.A.v.H., N.P., and N.Soranzo designed and coordinated the project.

J.W., R.T., and S.F. provided research resources.

F.P.C., N.P., B.S. and N.Soranzo supervised the study.

D.Z., M.T., B.S. and N.Soranzo wrote the manuscript.

All authors reviewed and approved the final manuscript.

## Consortia

### Genes & Health Research Team

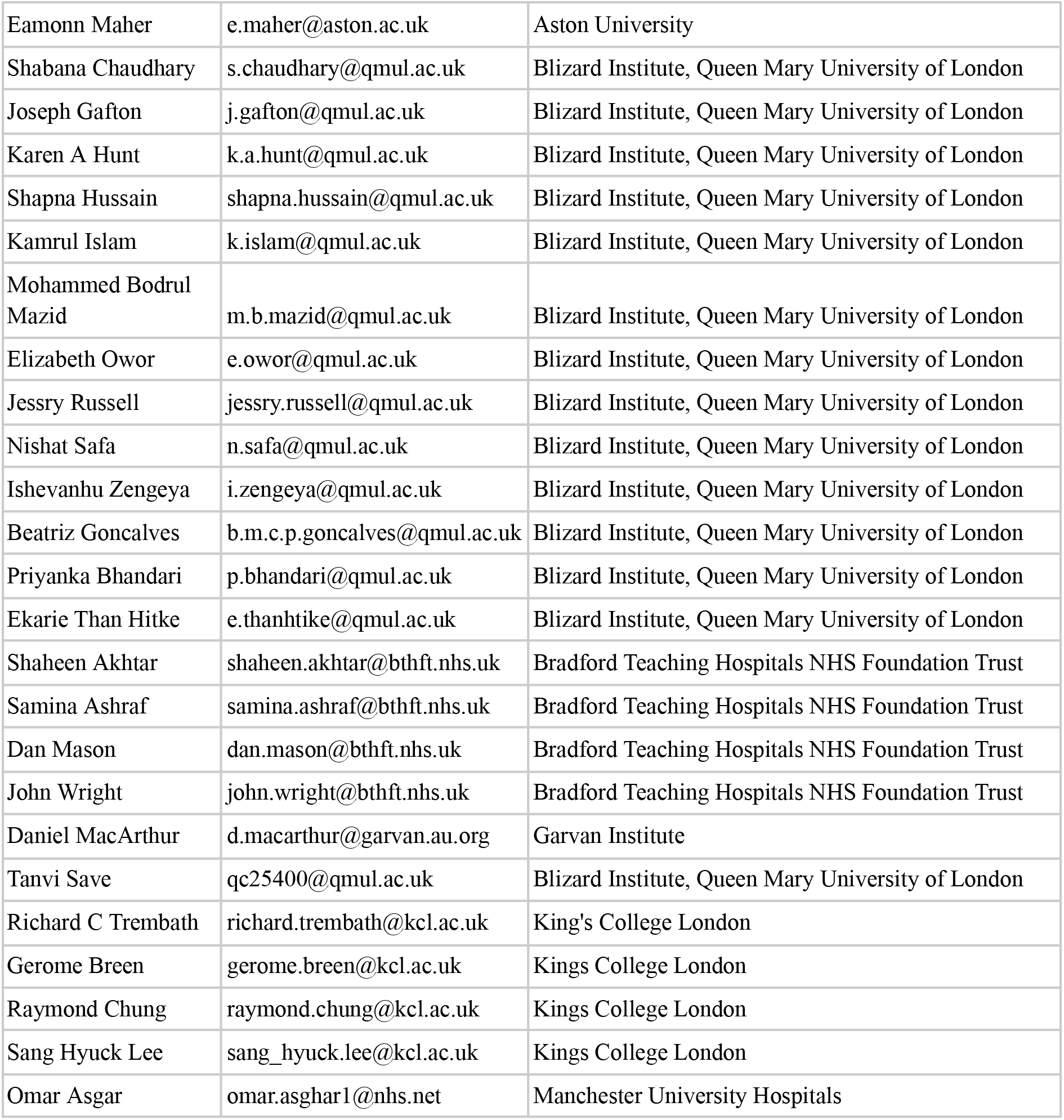
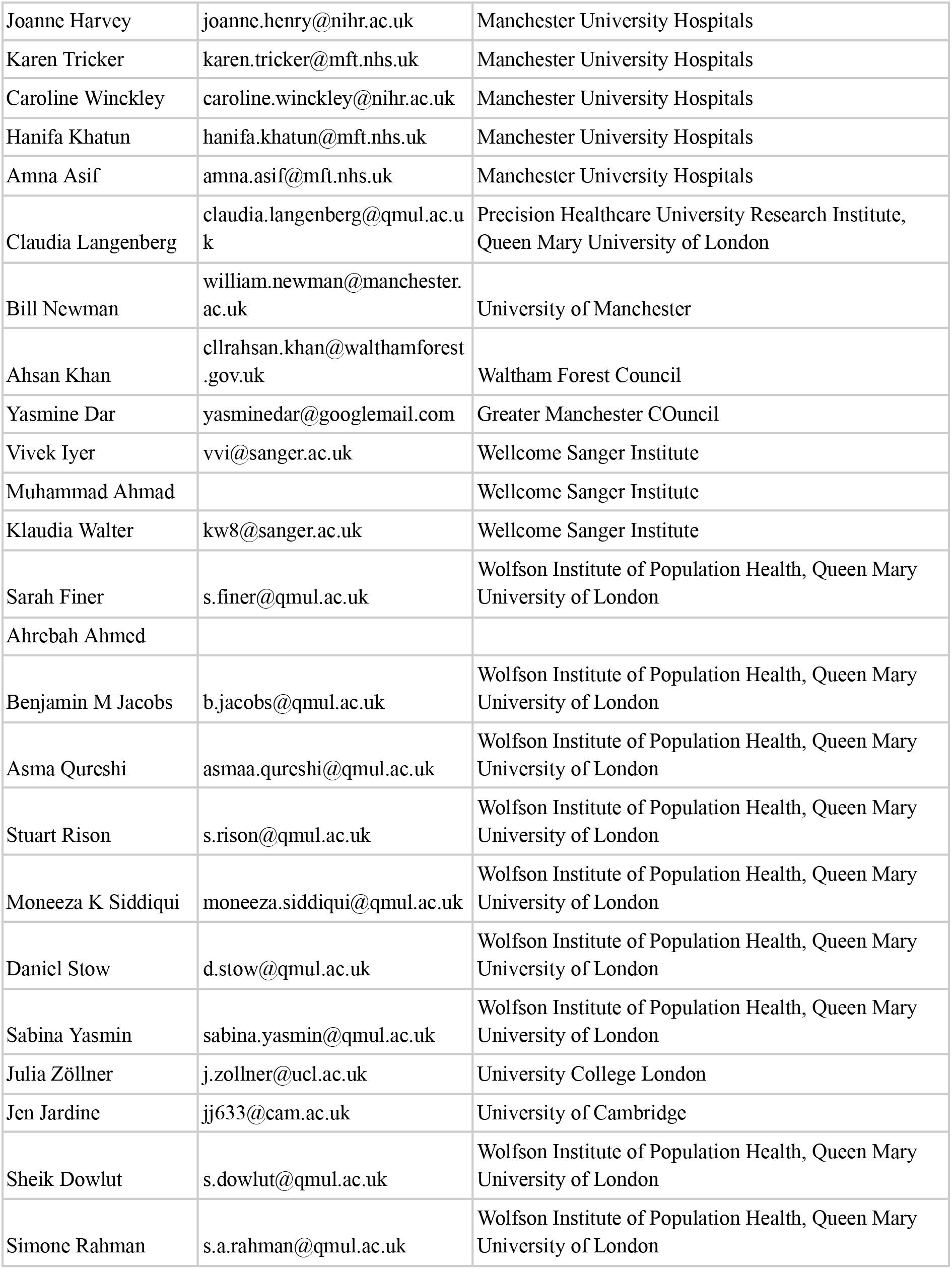

#### Data availability

The genotype and phenotype for the UKB cohort described in this study were obtained from the UK Biobank resource under approved application number 69938. These are available, together with raw and processed single-cell data, to bona fide researchers upon approved application to the UK Biobank Access Management System.

The health and genetic data for the G& H cohort used in this study were obtained from the Genes & Health resource. These are available, together with raw and processed single-cell data, to registered researchers via the secure Genes & Health Trusted Research Environment following approval by the cohort’s data access committee.

Single-cell complete blood counts (scCBCs) are available to bona fide researchers upon approved application to the UK Biobank Access Management System.

#### Code availability

Custom code utilised in this study is available via GitHub at https://github.com/DanielaZanotti/Zanotti_et_al_population_manuscript

